# Determining The Role Of Radiation Oncologist Demographic Factors On Segmentation Quality: Insights From A Crowd-Sourced Challenge Using Bayesian Estimation

**DOI:** 10.1101/2023.08.30.23294786

**Authors:** Kareem A. Wahid, Onur Sahin, Suprateek Kundu, Diana Lin, Anthony Alanis, Salik Tehami, Serageldin Kamel, Simon Duke, Michael V. Sherer, Mathis Rasmussen, Stine Korreman, David Fuentes, Michael Cislo, Benjamin E. Nelms, John P. Christodouleas, James D. Murphy, Abdallah S. R. Mohamed, Renjie He, Mohammed A. Naser, Erin F. Gillespie, Clifton D. Fuller

## Abstract

**BACKGROUND:** Medical image auto-segmentation is poised to revolutionize radiotherapy workflows. The quality of auto-segmentation training data, primarily derived from clinician observers, is of utmost importance. However, the factors influencing the quality of these clinician-derived segmentations have yet to be fully understood or quantified. Therefore, the purpose of this study was to determine the role of common observer demographic variables on quantitative segmentation performance.

**METHODS:** Organ at risk (OAR) and tumor volume segmentations provided by radiation oncologist observers from the Contouring Collaborative for Consensus in Radiation Oncology public dataset were utilized for this study. Segmentations were derived from five separate disease sites comprised of one patient case each: breast, sarcoma, head and neck (H&N), gynecologic (GYN), and gastrointestinal (GI). Segmentation quality was determined on a structure-by-structure basis by comparing the observer segmentations with an expert-derived consensus gold standard primarily using the Dice Similarity Coefficient (DSC); surface DSC was investigated as a secondary metric. Metrics were stratified into binary groups based on previously established structure-specific expert-derived interobserver variability (IOV) cutoffs. Generalized linear mixed-effects models using Markov chain Monte Carlo Bayesian estimation were used to investigate the association between demographic variables and the binarized segmentation quality for each disease site separately. Variables with a highest density interval excluding zero — loosely analogous to frequentist significance — were considered to substantially impact the outcome measure.

**RESULTS:** After filtering by practicing radiation oncologists, 574, 110, 452, 112, and 48 structure observations remained for the breast, sarcoma, H&N, GYN, and GI cases, respectively. The median percentage of observations that crossed the expert DSC IOV cutoff when stratified by structure type was 55% and 31% for OARs and tumor volumes, respectively. Bayesian regression analysis revealed tumor category had a substantial negative impact on binarized DSC for the breast (coefficient mean ± standard deviation: –0.97 ± 0.20), sarcoma (–1.04 ± 0.54), H&N (–1.00 ± 0.24), and GI (–2.95 ± 0.98) cases. There were no clear recurring relationships between segmentation quality and demographic variables across the cases, with most variables demonstrating large standard deviations and wide highest density intervals.

**CONCLUSION:** Our study highlights substantial uncertainty surrounding conventionally presumed factors influencing segmentation quality. Future studies should investigate additional demographic variables, more patients and imaging modalities, and alternative metrics of segmentation acceptability.

## Introduction

Segmentation (also termed contouring or delineation) of regions of interest (ROIs) on medical images is crucial for contemporary radiotherapy treatment planning ^1^. Importantly, accurate segmentation of organs at risk (OARs) and tumor-related structures are required to maximize radiotherapeutic efficacy while minimizing harmful side effects. Segmentation for radiotherapy treatment planning is often performed by highly trained clinicians, such as radiation oncologists. However, clinician-derived manual segmentation is a time– and labor-intensive task subject to significant inter-observer variation, thereby prompting the increasing development of artificial intelligence (AI)-based methods for auto-segmentation ^2^.

The contouring collaborative for consensus in radiation oncology (C3RO), a large-scale crowdsourcing challenge for radiotherapy segmentation, demonstrated that non-expert consensus ROI segmentations could quantitatively approximate expert consensus ROI segmentations in a variety of disease sites ^3^, thereby motivating the potential use of a large number of lower-quality segmentations in place of a small number of high-quality segmentations for AI auto-segmentation model training. Notably, segmentations were highly variable among the participants of C3RO, pointing to the existence of underlying factors associated with the resultant segmentation quality.

Despite AI advancements in auto-segmentation, human clinicians will likely be involved in the radiotherapy segmentation process for the foreseeable future, both as suppliers of ground truth segmentations for algorithmic training and as the final arbiters of auto-segmentation quality. Understanding the characteristics of radiation oncologists associated with superior segmentation performance is of utmost importance, as this knowledge can guide the training of future professionals, inform the design of auto-segmentation tools, and ultimately improve the quality of care provided to cancer patients. While some data do suggest that clinician experience in a particular disease site is associated with improved radiotherapy outcomes ^4–6^, no studies have directly examined underlying factors related to segmentation quality. Therefore, we aim to investigate how demographic factors of a large number of practicing radiation oncologists are associated with improved segmentation quality through a secondary analysis of the C3RO data across several disease sites.

## Methods

### Study participants and demographic variables

Participants in C3RO were categorized as recognized experts or non-experts. Recognized experts were identified by the C3RO organizers as board-certified physicians who participated in the development of national guidelines and/or contributed to extensive scholarly activities within a specific disease site. Non-experts were any participants not categorized as an expert for that disease site. For this study, non-expert participants from each separate disease site of the C3RO database, namely the breast, sarcoma, head and neck (H&N), gynecologic (GYN), and gastrointestinal (GI) cases were selected for the analysis. Greater details on the publicly available C3RO dataset can be found in the corresponding data descriptor ^7^. Self-reported demographic variables of interest from the participants were initially collected through an intake survey performed on REDCap ^8^. Demographic variables for this study included: practice location, self-identified gender, self-identified race, academic affiliation, primary practice type, number of radiation oncologist colleagues, presence of another radiation oncologist colleague on clinic day, and if the observer actively treated the disease site of interest. Additionally, a new demographic variable, years of practice, was calculated as the reported years since finishing residency minus the year C3RO data collection took place, i.e., 2022. **Table 1** shows the demographic variables in detail with corresponding descriptions and possible values. Before use in the analysis, non-expert participants were filtered out of the dataset if they were clinical residents (i.e., trainees) or non-clinicians. In other words, only currently practicing radiation oncologists were included in the analysis. Due to an imbalance between primary practice type groups, the primary practice description variable was converted to a binary format by grouping academic/university (academic) into one group and all others into a separate group (non-academic).

**Table 1.** Demographic variables examined in this study.

| Variable | Description |
| --- | --- |
| Practice location | Geographic location where participant actively practices. Binary variable with possible values of United States (US) or Non-US. |
| Gender | Self-identified gender. Binary variable with possible values of male or female. Original variable included non-binary as an option but was not selected by any participants. |
| Race | Self-identified race. Binary variable with possible values of white or non-white. |
| Academic Affiliation | Whether the participant actively holds an academic affiliation or not. Binary variable with possible values of yes or no. |
| Primary Practice Type | Best self-identified description of primary practice where participant actively practices. Categorical variables with values of, academic/university, non-academic hospital, private practice (solo or group), or other. Converted to a binary variable with possible values of academic or non-academic. |
| Number of Radiation Oncologist Colleagues | The total estimated number of radiation oncologist colleagues that work with the participant at their primary clinical site (excluding themselves). Continuous numerical variable with minimum value of 0. |
| Presence of Another Radiation Oncologist on Clinic Day | Whether there is at least one additional radiation oncologist when the participant is actively working in their clinic (on most days). Binary variable with possible values of yes or no. |
| Actively Treat Disease Site | Whether the participant actively treats the disease site under investigation, e.g., for the breast case do they actively treat breast patients in their clinic? Binary variable with possible values of yes or no. |
| Years of Practice | Number of self-reported years since completing residency (calculated relative to start of C3RO). Continuous numerical variable with minimum value of 0. |

### Segmentation evaluation

All ROIs from all disease sites in the C3RO dataset were used for this analysis. A complete list of ROIs and their corresponding abbreviations and descriptions can be found in **Appendix A**. For each non-expert ROI, we calculated segmentation quality by comparing the non-expert segmentation to the consensus of experts as derived using the Simultaneous Truth and Performance Level Estimation (STAPLE) algorithm ^9^ (**Figure 2**). The number of expert observers used for each ROI consensus segmentation can be found in **Appendix A**. The number of experts used to derive the consensus segmentation was variable depending on the ROI; more information can be found in the corresponding data descriptor ^7^. Though “experts” were subjectively determined in the original C3RO study, they demonstrated significantly improved interobserver variability compared to non-expert counterparts ^3^. Therefore, the expert STAPLE can be considered as a “gold standard” segmentation to be used for comparison purposes. We utilized existing Neuroimaging Informatics Technology Initiative (NIfTI) structure files for comparisons, which were originally converted from Digital Imaging and Communications in Medicine to NIfTI format using DICOMRTTool ^10^. The Dice similarity coefficient (DSC) was utilized as the main metric for comparison purposes due to its ubiquity in segmentation studies^1^. We also investigated a metric of surface similarity, the surface DSC (SDSC) for additional experiments; tolerance values for each ROI were determined from the pairwise average surface distance of the expert segmentations which can be found in the C3RO data descriptor ^7^. Metrics were calculated using the surface-distances Python package v. 0.1 ^11^ and in-house Python code (Python v. 3.9.0).

**Figure 1.**
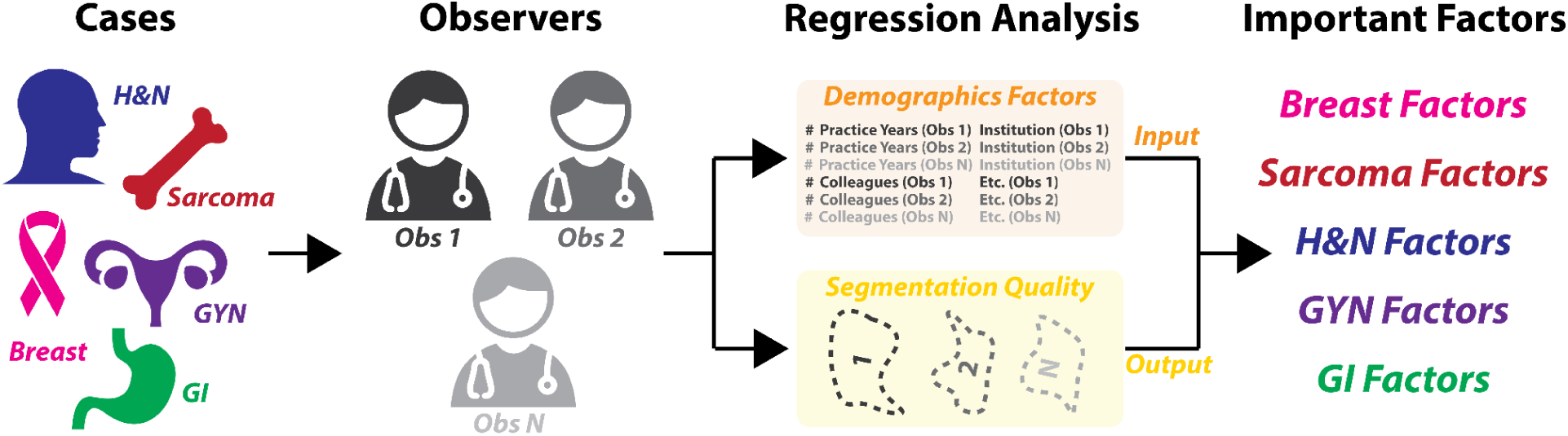
Overview of study. Five cases from different disease sites (breast, sarcoma, head and neck [H&N], gynecologic [GYN], and gastrointestinal [GI]) were investigated. Radiation oncologist observers segmented organs at risk and tumor-related structures from these cases, whereupon Bayesian regression analysis was performed to determine the relationship between underlying demographic factors and segmentation quality.

**Figure 2.**
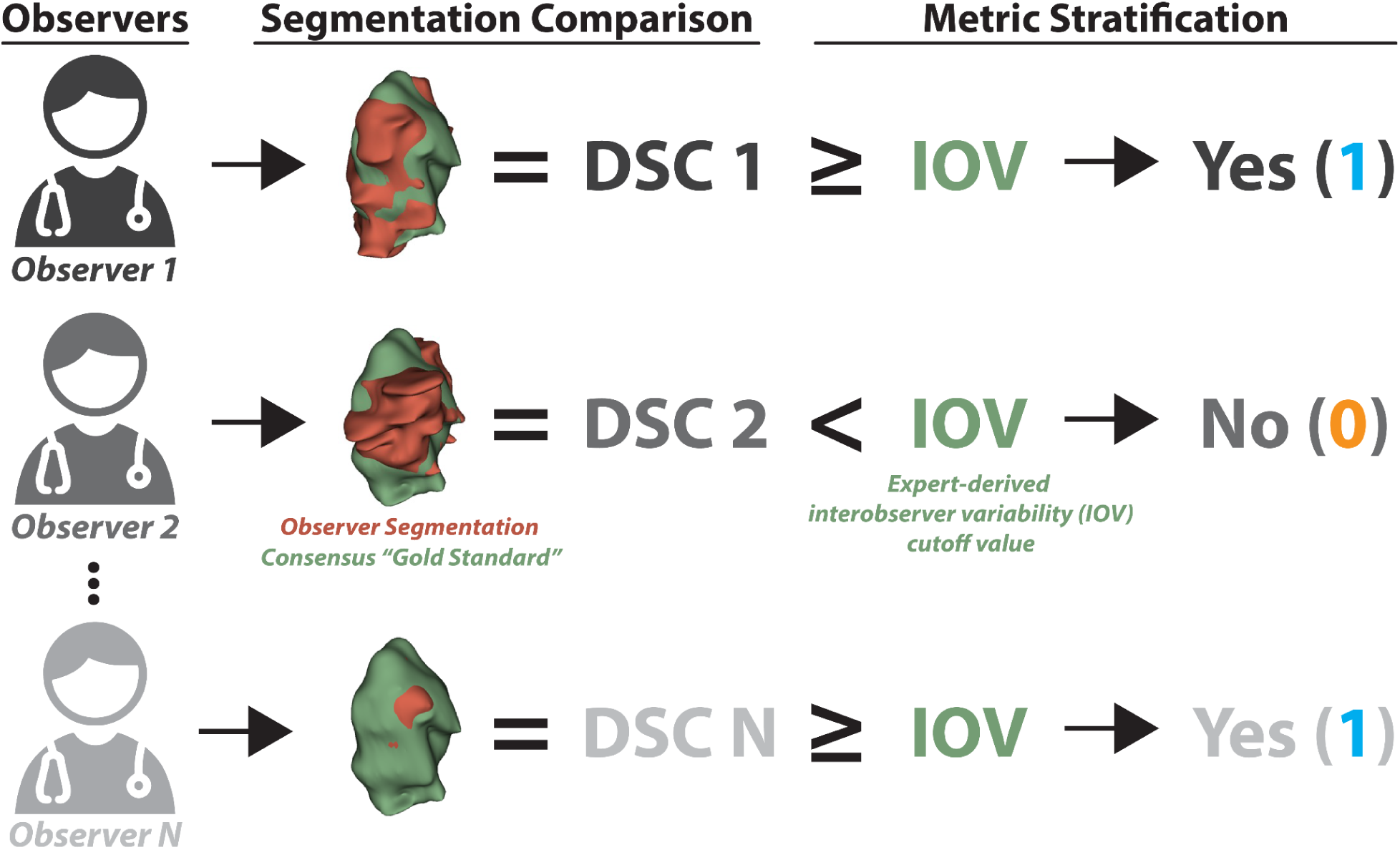
Derivation of binarized structure segmentation quality for each observer. Each observer could segment multiple structures, i.e., organs at risk and tumor volumes. Observer segmentations (red volume) were compared to a “gold standard” derived from a consensus segmentation of experts (green volume) using the Dice similarity coefficient (DSC). Segmentation metrics were then stratified into being greater than or equal to (yes – 1) or below (no – 0) a previously derived expert-derived interobserver variability (IOV) cutoff value for that particular region of interest. In this example, the primary gross tumor volume structure for the head and neck case is shown. A similar process was used to derive binarized values for surface DSC.

In order to ensure metrics were comparable across ROIs, metrics were stratified into binary groups based on previously established ROI-specific expert-derived interobserver variability (IOV) cutoffs ^7^. Namely if the metric for a given ROI was greater than or equal to the ROI-specific expert IOV, it was classified as a 1, while if it was less than the expert IOV it was classified as a 0 (**Figure 2**). For example, if an observer scored a DSC of 0.95 for the ROI “heart” whose expert IOV was 0.9, the binarized DSC value would be 1. Finally, for each ROI, we calculated the percentage of observers that were able to cross the expert IOV cutoff.

### Bayesian regression analysis

Due to the repeated measures nature of our study, generalized linear mixed effects models with Bayesian estimation were utilized to investigate the relationship between demographic variables and binarized segmentation quality metrics for each disease site separately. A Bernoulli logistic model was implemented due to the binary nature of our outcome variable. The stratified binary segmentation quality metric (i.e., IOV thresholded metric of non-expert relative to expert STAPLE) acted as the dependent variable for the models. The key independent variables (i.e., fixed effects) were practice location, primary practice type, number of radiation oncologist colleagues, presence of another radiation oncologist on clinic day, actively treated the disease site, and years of practice. Notably, exploratory correlative analysis (**Appendix B**) revealed high relative correlation between academic affiliation and primary practice type; therefore academic affiliation, the less initially granular of the two variables, was not included as a covariate in this work to facilitate model parsimony. An additional binary categorical variable, ROI type, was added as an additional independent variable to indicate if the ROI was an OAR or tumor volume. Additionally, models were corrected for self-identified gender and self-identified race by including them as independent variables in the models. A random intercept was used in the models to account for the various observers who could segment multiple structures on the same image (e.g., multiple OARs and tumor volumes). In other words, independent observers were treated as groups for the mixed effect models. Any empty values for numerical variables, namely number of radiation oncologist colleagues and years of practice, were imputed to the median value relative to the total number of observations for that disease site. Finally, numerical variables were Z-score normalized within each separate disease site to facilitate model convergence and direct comparison of coefficient values.

The Python package Bambi v. 0.12.0 ^12^, which is built on top of the robust Markov chain Monte Carlo (MCMC) library PyMC3 ^13^, was utilized for all regression analysis. For each disease site (breast, sarcoma, H&N, GYN, or GI), the regression formula was defined as:

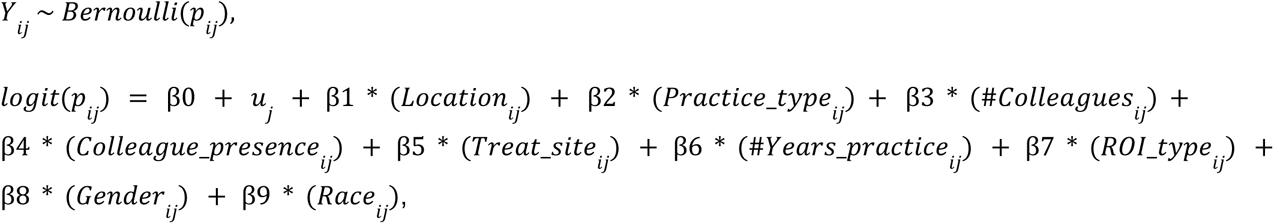

Where, *y_ij_* is the dependent variable (either binarized DSC or SDSC) for observation *i* nested within observer *j* which follows a Bernoulli distribution with success probability *p_ij_*; *logit*(*p_ij_*) is the log-odds of the success probability; β0 is the overall intercept; *u_j_* is the random intercept for observer *j*; β1, …, β9 are the fixed effect coefficients for the predictors, which also have interpretations in terms of odds ratios under a logistic regression framework. Number of colleagues and number of years of practice were numerical variables, while all other demographic variables were binary categorical variables.

For each MCMC Bayesian regression model, 10,000 samples were drawn from 4 chains with a tuning set of 1500 iterations for a total of 46,000 samples drawn for each model. A random state was set to ensure a reproducible model fitting procedure. Weakly informative priors as determined by the Bambi package were intelligently generated for all model terms by loosely scaling them to the observed data ^12^. Computations were performed across 6 cores of an Intel® Core™ i7-8700 Processor. MCMC Bayesian regression model computation took between 1-4 hours for each model.

The ArviZ v. 0.15.1 ^14^ Python library was used to derive summary data for the posterior distribution. Point estimates (posterior means) and assessments of uncertainty (posterior standard deviation) were calculated for each variable. Additionally, the 89% highest density interval (HDI) — also referred to as the minimum width Bayesian credible interval — was calculated; a value of 89% was selected as suggested by recent literature ^15,16^. ArviZ computes the HDI using an empirical method based on the sorted posterior samples; additional information on ArviZ calculations can be found in the corresponding documentation and source code ^14^. When an HDI does not contain zero, it suggests that the true value of the parameter is either entirely positive or negative. Therefore, demographic variables for which the HDI did not include zero were considered to have a substantial impact on the outcome measure of interest and could be interpreted as loosely analogous to the frequentist notion of statistical significance.

### Data and code availability

All C3RO data, including the original demographic factors and segmentation data are available on Figshare (DOI = doi.org/10.6084/m9.figshare.21074182). All Python code used for generating and analyzing the data for this study is available on GitHub (URL = https://github.com/kwahid/C3RO_demographics_analysis). Corresponding newly created data and spreadsheets generated for this study can also be found on Figshare (DOI = doi.org/10.6084/m9.figshare.24021591).

## Results

### Study participants

Flow diagrams of the number of structures and number of clinician observers investigated for each disease site are shown in **Figure 3**. After filtering out structures from non-eligible observers, 574, 110, 452, 112, and 48 ROI structure observations from practicing radiation oncologist observers remained for the analysis for the breast, sarcoma, H&N, GYN, and GI cases, respectively.

**Figure 3.**
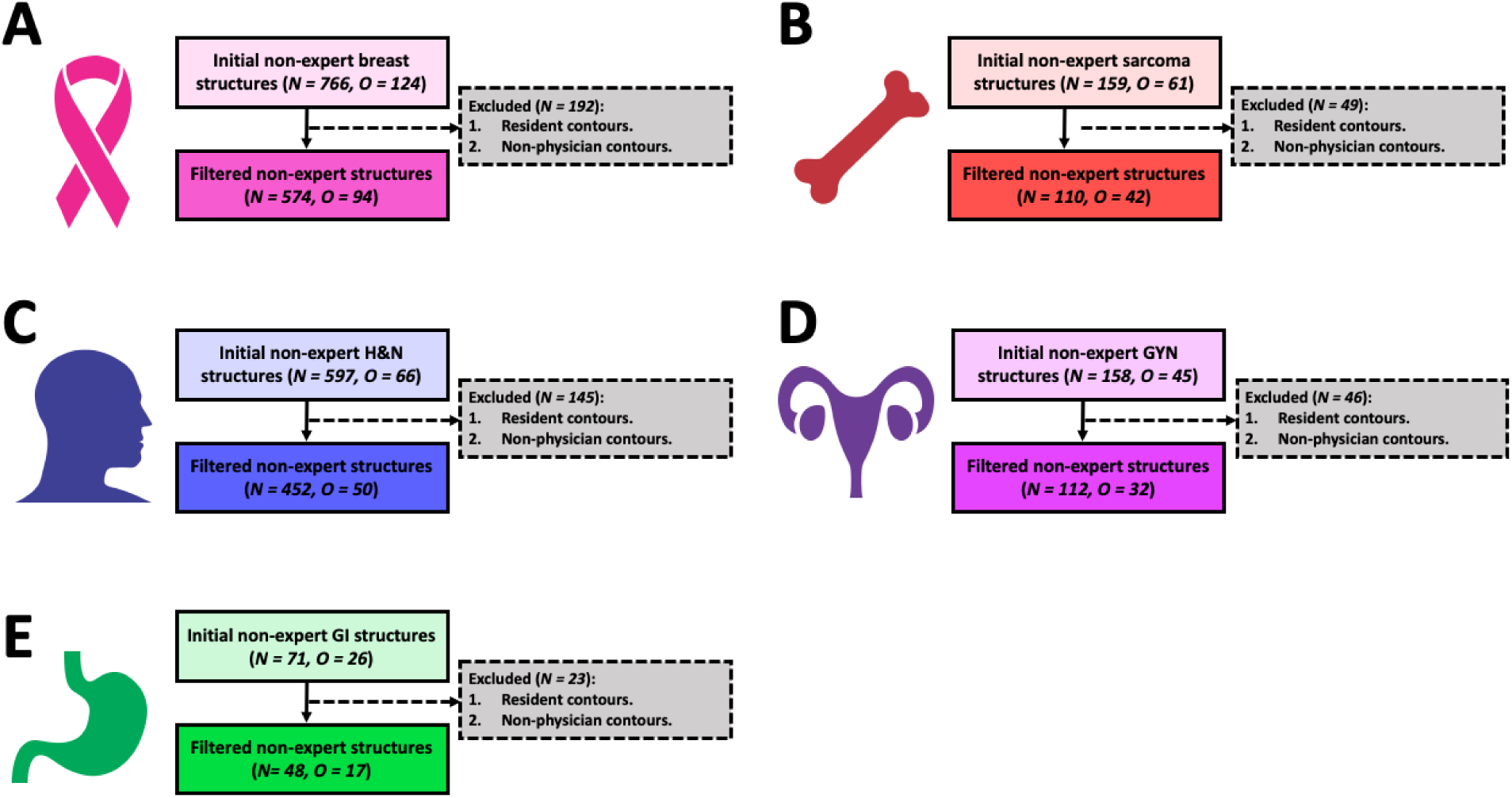
Flow diagrams showing the final number of structures evaluated for each disease site. Breast, sarcoma, H&N, GYN, and GI cases are shown in panels **(A)**, **(B)**, **(C)**, **(D)**, and **(E)**, respectively. Abbreviations: H&N = head and neck, GYN = gynecologic, GI = gastrointestinal, N = number of non-expert structure segmentations, O = number of unique non-expert observers.

### Individual observer performance

**Figure 4** shows the DSC scores for each observer relative to the expert consensus segmentation stratified by ROI; the percentage of observations that were able to cross the expert IOV cutoff are also shown. The highest percentages per case were BrachialPlex_L (82%), Genitals (44%), Glnd_Submand_L (76%), GTV_n (70%), and Bag_Bowel (73%) for breast, sarcoma, H&N, GYN, and GI respectively. The lowest percentages per case were CTV_IMN (36%), CTV (18%), GTVn (24%), CTVn_4500 (26%), and CTV_4500 (29%) for breast, sarcoma, H&N, GYN, and GI respectively. Aggregated median percentage values when stratified by ROI type were 55% (interquartile range [IQR] = 35%) and 31% (IQR = 15%) for OARs and tumor volumes, respectively. Analogous bar plots using SDSC as a metric are shown in **Appendix C**. SDSC values mirrored DSC values for most ROIs; aggregated SDSC median percentage values when stratified by ROI type were 36% (IQR = 32%) and 30% (IQR = 30%) for OARs and tumor volumes, respectively.

**Figure 4.**
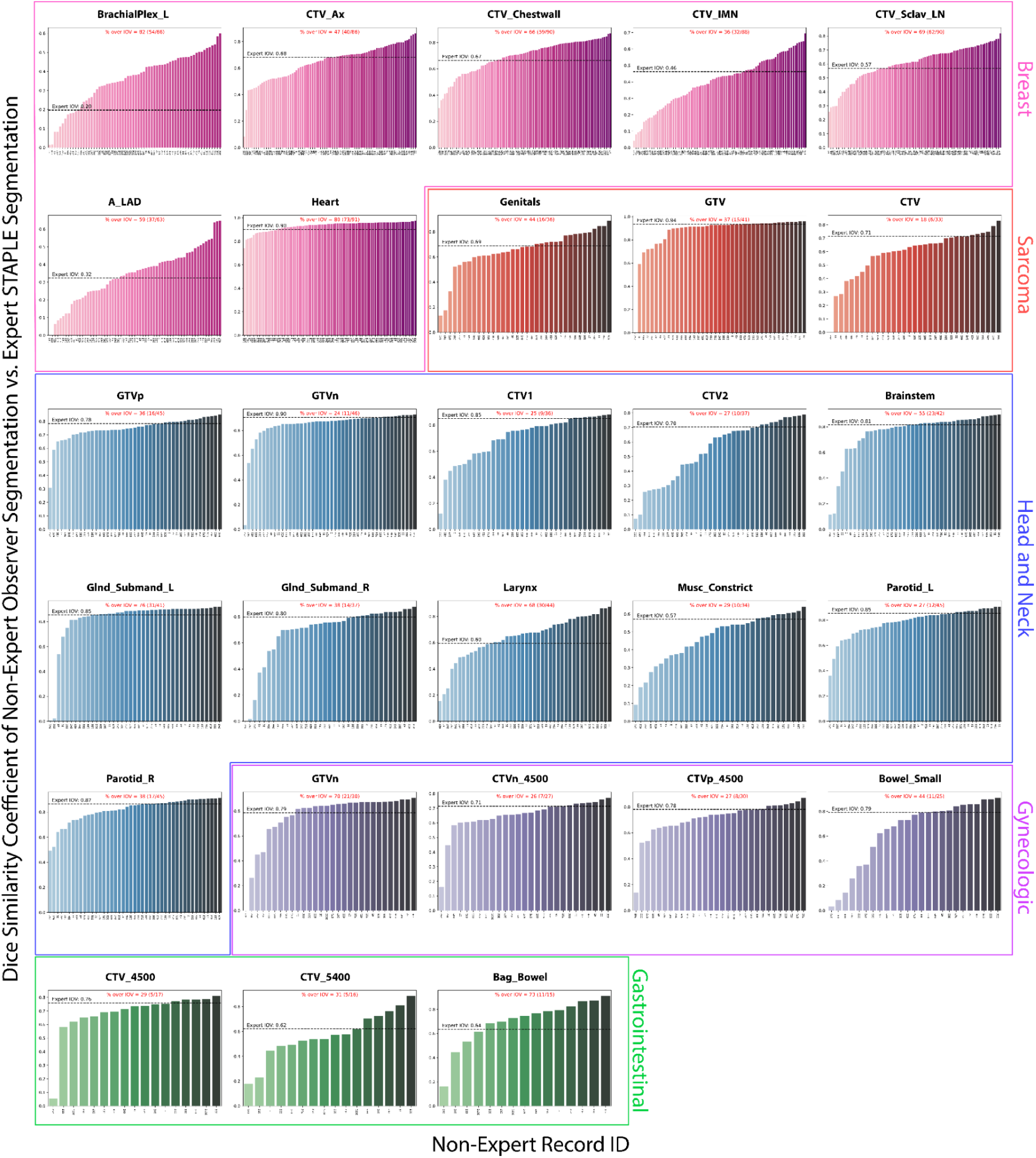
Barplots of individual observer segmentation performance vs. gold standard. Pink, red, blue, purple, and green plots correspond to breast, sarcoma, head and neck, gynecologic, and gastrointestinal regions of interest, respectively. The gold standard segmentation is the consensus segmentation of all experts as derived from the Simultaneous Truth and Performance Level Estimation algorithm. Black dotted lines indicate median expert interobserver dice similarity coefficient (DSC) for a corresponding region of interest. The percentage of observers that crossed the expert interobserver variability (IOV) cutoff is shown in red above each plot.

### Bayesian regression models

Mixed effects regression results using binarized DSC and binarized SDSC as the outcome variables are shown in **Table 2** and **Table 3**, respectively. For the breast case, tumor category for both DSC (mean±SD: –0.97±0.20) and SDSC (–1.24±0.20) had HDIs that excluded zero. For the sarcoma case, tumor category for both DSC (–1.04±0.54) and SDSC (–2.74±0.81) had HDIs that excluded zero. For the H&N case, the DSC tumor category (–1.00±0.24) and DSC white racial self-identification (0.66±0.41) had HDIs that excluded zero. For the GYN case, only SDSC academic practice type (–1.30±0.79) had an HDI that excluded zero. For the GI case, DSC tumor category (–2.95±0.98) and DSC colleague presence (2.21±1.40) had HDIs that excluded zero. Model convergence parameters estimated for each variable are presented in **Appendix D**.

**Table 2.** Generalized linear mixed-effects models with Bayesian estimation results using binarized Dice similarity coefficient as the outcome variable. Model coefficient values are shown for each variable. Reference variable for categorical variables are shown in brackets next to variable name. Sign value in posterior mean indicates positive or negative correlation of variable with outcome. Posterior standard deviation (SD) indicates uncertainty around posterior mean. 89% highest density interval (HDI) is shown in parenthesis after posterior mean. * Bolded variables indicate HDI does not contain zero and is considered to have a substantial impact on the outcome measure of interest.

|  | Breast |  | Sarcoma |  | Head and Neck |  | Gynecologic |  | Gastrointestinal |  |
| --- | --- | --- | --- | --- | --- | --- | --- | --- | --- | --- |
| Variables | Mean (HDI) | SD | Mean (HDI) | SD | Mean (HDI) | SD | Mean (HDI) | SD | Mean (HDI) | SD |
| Intercept | 0.90 (0.08,1.69) | 0.51 | 0.76 (-1.30,2.93) | 1.35 | -0.46 (-1.85,0.87) | 0.86 | -0.10 (-2.02,1.78) | 1.21 | -3.28 (-7.34,1.08) | 2.67 |
| ROI category [Tumor] | **-0.97 (-1.29,-0.65)\*** | 0.20 | **-1.04 (-1.91,-0.19)\*** | 0.54 | **-1.00 (-1.39,-0.62)\*** | 0.24 | -0.09 (-0.92,0.72) | 0.51 | **-2.95 (-4.44,-1.36)\*** | 0.98 |
| Location [US] | -0.54 (-1.15,0.06) | 0.38 | 0.55 (-1.16,2.21) | 1.07 | 0.07 (-0.92,1.06) | 0.62 | 0.08 (-1.52,1.66) | 1.01 | 4.24 (-0.69,9.36) | 3.22 |
| Gender [Female] | -0.10 (-0.51,0.31) | 0.26 | 0.66 (-0.60,1.91) | 0.80 | -0.26 (-1.01,0.47) | 0.47 | -0.13 (-1.15,0.88) | 0.64 | 0.55 (-1.25,2.35) | 1.14 |
| Years of practice | -0.06 (-0.25,0.13) | 0.12 | -0.44 (-1.07,0.18) | 0.41 | -0.23 (-0.56,0.11) | 0.21 | -0.25 (-0.78,0.24) | 0.33 | -0.33 (-1.49,0.83) | 0.74 |
| Practice type [Academic] | -0.37 (-0.74,0.02) | 0.24 | -0.01 (-1.24,1.21) | 0.79 | 0.13 (-0.48,0.75) | 0.39 | -0.37 (-1.34,0.59) | 0.62 | -1.37 (-3.87,1.12) | 1.58 |
| # of Colleagues | -0.02 (-0.22,0.18) | 0.12 | -0.15 (-0.82,0.53) | 0.43 | 0.22 (-0.15,0.58) | 0.23 | -0.13 (-0.61,0.36) | 0.31 | 0.01 (-1.51,1.59) | 0.98 |
| Colleague presence [Yes] | 0.28 (-0.21,0.77) | 0.31 | -1.13 (-2.60,0.37) | 0.95 | 0.24 (-0.62,1.07) | 0.53 | 0.23 (-1.06,1.57) | 0.83 | **2.21 (0.03,4.43)\*** | 1.40 |
| Race [White] | -0.30 (-0.68,0.09) | 0.24 | -0.77 (-2.06,0.53) | 0.83 | **0.66 (0.02,1.33)\*** | 0.41 | 0.47 (-0.47,1.36) | 0.58 | 0.89 (-0.91,2.70) | 1.15 |
| Treat disease site [Yes] | 0.53 (-0.14,1.20) | 0.42 | -0.39 (-1.62,0.84) | 0.82 | -0.13 (-1.33,1.02) | 0.74 | -0.42 (-1.76,0.92) | 0.85 | 3.02 (-0.34,6.26) | 2.09 |
| Random effect variance | 0.57 (0.27,0.90) | 0.19 | 1.44 (0.18,2.47) | 0.74 | 1.03 (0.67,1.39) | 0.23 | 0.73 (0.00,1.31) | 0.46 | 0.78 (0.00,1.58) | 0.64 |

**Table 3.**
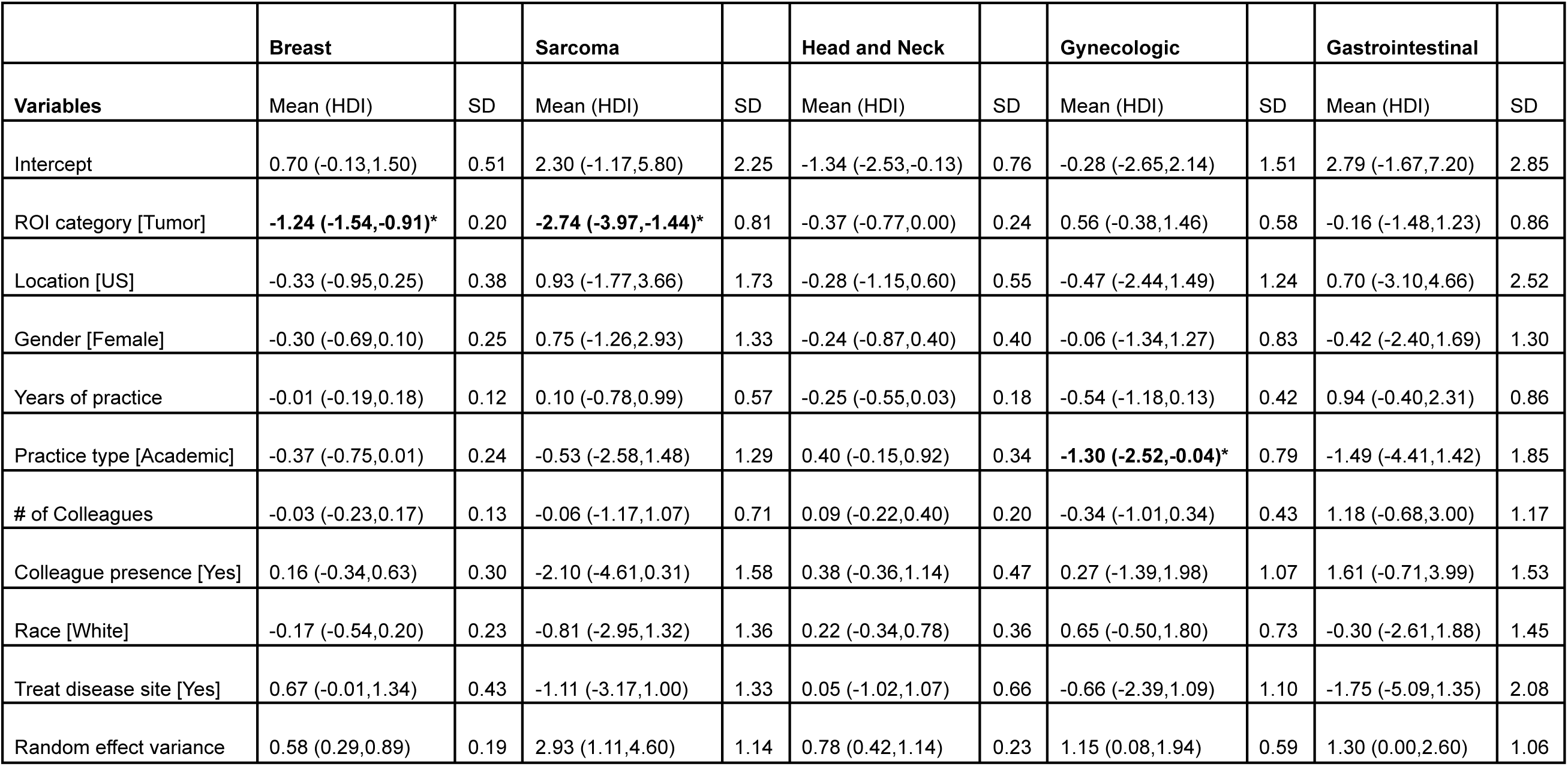
Generalized linear mixed-effects models with Bayesian estimation results using binarized surface Dice similarity coefficient as the outcome variable. Model coefficient values are shown for each variable. Reference variable for categorical variables are shown in brackets next to variable name. Sign value in posterior mean indicates positive or negative correlation of variable with outcome. Posterior standard deviation (SD) indicates uncertainty around posterior mean. 89% highest density interval (HDI) is shown in parenthesis after posterior mean. * Bolded variables indicate HDI does not contain zero and is considered to have a substantial impact on the outcome measure of interest.

|  | Breast |  | Sarcoma |  | Head and Neck |  | Gynecologic |  | Gastrointestinal |  |
| --- | --- | --- | --- | --- | --- | --- | --- | --- | --- | --- |
| Variables | Mean (HDI) | SD | Mean (HDI) | SD | Mean (HDI) | SD | Mean (HDI) | SD | Mean (HDI) | SD |
| Intercept | 0.70 (-0.13,1.50) | 0.51 | 2.30 (-1.17,5.80) | 2.25 | -1.34 (-2.53,-0.13) | 0.76 | -0.28 (-2.65,2.14) | 1.51 | 2.79 (-1.67,7.20) | 2.85 |
| ROI category [Tumor] | **-1.24 (-1.54,-0.91)\*** | 0.20 | **-2.74 (-3.97,-1.44)\*** | 0.81 | -0.37 (-0.77,0.00) | 0.24 | 0.56 (-0.38,1.46) | 0.58 | -0.16 (-1.48,1.23) | 0.86 |
| Location [US] | -0.33 (-0.95,0.25) | 0.38 | 0.93 (-1.77,3.66) | 1.73 | -0.28 (-1.15,0.60) | 0.55 | -0.47 (-2.44,1.49) | 1.24 | 0.70 (-3.10,4.66) | 2.52 |
| Gender [Female] | -0.30 (-0.69,0.10) | 0.25 | 0.75 (-1.26,2.93) | 1.33 | -0.24 (-0.87,0.40) | 0.40 | -0.06 (-1.34,1.27) | 0.83 | -0.42 (-2.40,1.69) | 1.30 |
| Years of practice | -0.01 (-0.19,0.18) | 0.12 | 0.10 (-0.78,0.99) | 0.57 | -0.25 (-0.55,0.03) | 0.18 | -0.54 (-1.18,0.13) | 0.42 | 0.94 (-0.40,2.31) | 0.86 |
| Practice type [Academic] | -0.37 (-0.75,0.01) | 0.24 | -0.53 (-2.58,1.48) | 1.29 | 0.40 (-0.15,0.92) | 0.34 | **-1.30 (-2.52,-0.04)\*** | 0.79 | -1.49 (-4.41,1.42) | 1.85 |
| # of Colleagues | -0.03 (-0.23,0.17) | 0.13 | -0.06 (-1.17,1.07) | 0.71 | 0.09 (-0.22,0.40) | 0.20 | -0.34 (-1.01,0.34) | 0.43 | 1.18 (-0.68,3.00) | 1.17 |
| Colleague presence [Yes] | 0.16 (-0.34,0.63) | 0.30 | -2.10 (-4.61,0.31) | 1.58 | 0.38 (-0.36,1.14) | 0.47 | 0.27 (-1.39,1.98) | 1.07 | 1.61 (-0.71,3.99) | 1.53 |
| Race [White] | -0.17 (-0.54,0.20) | 0.23 | -0.81 (-2.95,1.32) | 1.36 | 0.22 (-0.34,0.78) | 0.36 | 0.65 (-0.50,1.80) | 0.73 | -0.30 (-2.61,1.88) | 1.45 |
| Treat disease site [Yes] | 0.67 (-0.01,1.34) | 0.43 | -1.11 (-3.17,1.00) | 1.33 | 0.05 (-1.02,1.07) | 0.66 | -0.66 (-2.39,1.09) | 1.10 | -1.75 (-5.09,1.35) | 2.08 |
| Random effect variance | 0.58 (0.29,0.89) | 0.19 | 2.93 (1.11,4.60) | 1.14 | 0.78 (0.42,1.14) | 0.23 | 1.15 (0.08,1.94) | 0.59 | 1.30 (0.00,2.60) | 1.06 |

## Discussion

Auto-segmentation, primarily based on deep learning, is primed to play a major role in radiotherapy workflows of the future. It is well established that training these auto-segmentation algorithms requires high-quality curated segmentation data derived from clinician observers. Thus far, the underlying factors of what makes a “good” clinician segmenter in a quantitative sense are unknown. In this study, we utilized generalized linear mixed effects models with Bayesian estimation to determine the relationship of practicing radiation oncologist demographic variables with radiotherapy-related segmentation quality as derived from quantitative metrics. Our study is the first to investigate the role of demographic variables on segmentation quality using a large set of clinician observers and OAR/tumor structures.

To date, there are limited objective and standardized measures to evaluate radiotherapy-related segmentation quality. Nissen et al. recently proposed the utilization of the Jaccard Index, a close analog to the DSC, for longitudinal quantitative radiation oncology resident evaluation ^17^. However, the inherent quality discerned from these metrics in their raw numerical form often varies based on the specific ROI. For example, a DSC of 0.80 for a particularly “simple” OAR may be less desirable than a DSC of 0.80 for a particularly “difficult” tumor volume, and thus raw metrics may not be immediately clinically useful. However, stratification of evaluation metrics, as we have performed in our study, allows for ROI-specific thresholds that act as rough measures of clinical acceptability. Notably, our ROI-specific thresholds are derived from “gold standard” measurements provided by recognized experts within particular disease sites, which were established to have significantly improved segmentation consistency compared to non-experts in previous work ^3^. When stratified by previously defined expert IOV cutoffs, the ROIs with the lowest percentage of observers that were able to cross cutoffs were often tumor volumes. This is consistent with the generally held notion that tumor volumes, which often require domain-specific knowledge, are inherently more difficult to segment than OARs and are prone to high variation ^18,19^; these results are echoed in our previous work ^3^. Consistent with the aforementioned results, Bayesian regression analysis demonstrated that tumor-related ROI categories adversely affected segmentation performance. This impact was evident for most disease sites using DSC and several sites using SDSC.

Interestingly, results were inconsistent and mostly non-substantial for the majority of demographic variables across disease sites. However, the extensive uncertainties associated with the various demographic variables, even those that excluded zero in their HDIs, are clearly illustrated by correspondingly large standard deviation values and HDI widths. Historically, greater institutional support has been perceived to be important for radiotherapy quality ^20^.

Therefore, our mostly negative results for proxy variables intuitively linked to greater institutional resource support, such as academic practice and factors related to radiation oncologist colleagues, are particularly surprising. While existing literature regarding observer demographic impact on radiotherapy-related tasks is sparse, it warrants mentioning that one of the few studies in this area found no significant relationship between demographic factors and the resultant quality of radiotherapy plans ^21^. Outside of radiotherapy applications, a similar study that focused on crowdsourcing radiologic annotations of lung diseases demonstrated no impact of observer demographics on segmentation quality ^22^. These studies echo our mostly null results.

While most of the investigated demographic variables were non-substantial with large degrees of uncertainties, there were a few notable results that we believe warrant further discussion. Academic practice in the GYN case was substantially negatively associated with SDSC performance; a non-substantial negative association was echoed in most of the disease sites. This could imply, perhaps contrary to common assumptions, that community clinicians produce segmentations more closely aligned with our gold standard and, presumably, more consistent with contouring guidelines. Moreover, white racial self-identification was substantially positively associated with DSC performance in the H&N case, which exhibited conflicting relationships in other disease sites. It’s crucial to emphasize that the association between racial self-identification — a complex social construct which has been drastically simplified in this binary variable — and segmentation performance likely reflects broader institutional or regional conformance to contouring guidelines, rather than a reductive racial skill disparity. Notably, US and European organizations, which would have overrepresentation of white racial self-identification, have the largest proportion of contouring guideline endorsements ^23^. The heterogeneity within C3RO’s categorization of non-U.S. observers may have confounded these relationships in our analysis. Additionally, the presence of a radiation oncologist colleague was substantially positively associated with DSC performance in the GI case; this positive relationship seemed to hold for most disease sites. These results suggest that clinicians who likely participate in consensus decision-making (e.g., peer-review) tend to create segmentations closer to our gold standard, and thus are likely to adhere to guidelines. Perplexingly, years of practice was found to have a consistently negative (though non-substantial) impact on DSC performance across the various disease sites. This may be because recent clinician graduates are more likely to adhere to contouring guidelines. Finally, our study did not show that treatment of a particular disease site was substantially associated with superior segmentation quality; in fact, it often demonstrated a negative correlation. This seemingly challenges previous findings highlighting the significant role of clinician experience in treatment quality ^4–6^. However, the variable did not assess treatment frequency for the specific site, thereby potentially introducing heterogeneity in its interpretation and ultimately diminishing its utility.

Our study is not without limitations. Firstly, we relied on an existing dataset with inherent constraints. While boasting an unprecedented number of individual radiation oncologist observers (>200), C3RO only principally utilized a single imaging modality (computed tomography) from one representative patient per disease site. While this provides a dedicated reference standard, the demographic relationships could change depending on a variety of underlying patient-related factors (e.g., disease complexity, image modality availability).

Moreover, the C3RO intake survey — from which demographic variables for our models were derived — was self-reported and requested limited demographic information. For example, direct indicators of treatment volume, which have been shown in previous studies to be strongly correlated to patient outcomes in several disease sites ^4^ were not collected due to the high potential for recall bias. Similarly, variables related to the routine use of contour guidelines in clinician workflows would have also likely been highly informative but were not collected.

Secondly, we have relied exclusively on conventional geometry-based metrics of segmentation quality (e.g., DSC, SDSC), which have been noted to have significant flaws in the assessment of radiotherapy-related structures ^1^. Future studies should investigate metrics more closely tied to relevant patient outcomes, such as dose-volume histogram measures. On a related note, how to best define segmentation quality in a quantitative manner, and subsequently how to improve it, remains an open question. We hope to mitigate some of these issues by binarizing our outcome segmentation quality variable, and thus calibrating the value relative to a gold standard baseline (i.e., expert interobserver variability). We fully acknowledge that this methodology has flaws, principally in that “edge cases” may be unfairly penalized or rewarded. However, in the context of educational tools, we believe these methods may be useful for initial quantitative assessment; we might recommend combining cutoff values of complementary metrics, such as DSC and SDSC, to gauge segmentation initial “passability”. Naturally, further refinement of metric utilization will likely be necessary and be context-specific, such as prioritizing the minimization of false negatives for tumor-related volumes. A final limitation of our study lies in our reliance on weakly informative priors for our Bayesian analysis, primarily due to insufficient existing data to extract meaningful priors from this under-researched topic. Nevertheless, our current data can serve as valuable priors for future Bayesian analyses.

## Conclusion

In summary, we utilized an extensive number of practicing radiation oncologist observers in several disease sites to probe trends between common demographic variables and segmentation quality using generalized linear mixed effects models with Bayesian estimation. Tumor-related structures were, as expected, more difficult to segment than organs at risk. However, results for demographic factors were mixed and exhibited high uncertainty as evident by large posterior standard deviations and wide highest density intervals. Surprisingly, there were no obvious recurring relationships for conventionally presumed factors influencing segmentation quality (e.g., measures of greater institutional resource support or actively treating the disease site). While stark variations in quantitative performance among observers compared to our gold standard segmentations can be observed, it is still unclear if and how demographic factors influence segmentation quality. Given the anticipated scenario that auto-segmentation algorithms will still require humans in the loop in some capacity, these factors are still likely important to understand. By tapping into a large public dataset that supports repeat analyses and data pooling, our study lays the foundation for further investigations into the factors that influence human segmentation performance. Future studies should investigate a greater number of demographic variables (e.g., direct indicators of treatment volume), a greater number of patients and imaging modalities, and alternative metrics of segmentation acceptability (e.g., dosimetric indicators).

## Data Availability

Corresponding newly created data and spreadsheets generated for this study can also be found on Figshare (DOI = doi.org/10.6084/m9.figshare.24021591).

## Acknowledgments

The authors thank Dr. Charles R. Thomas Jr. for helpful comments and discussions.

## Appendix A: Additional C3RO descriptive information

**Table A1.**
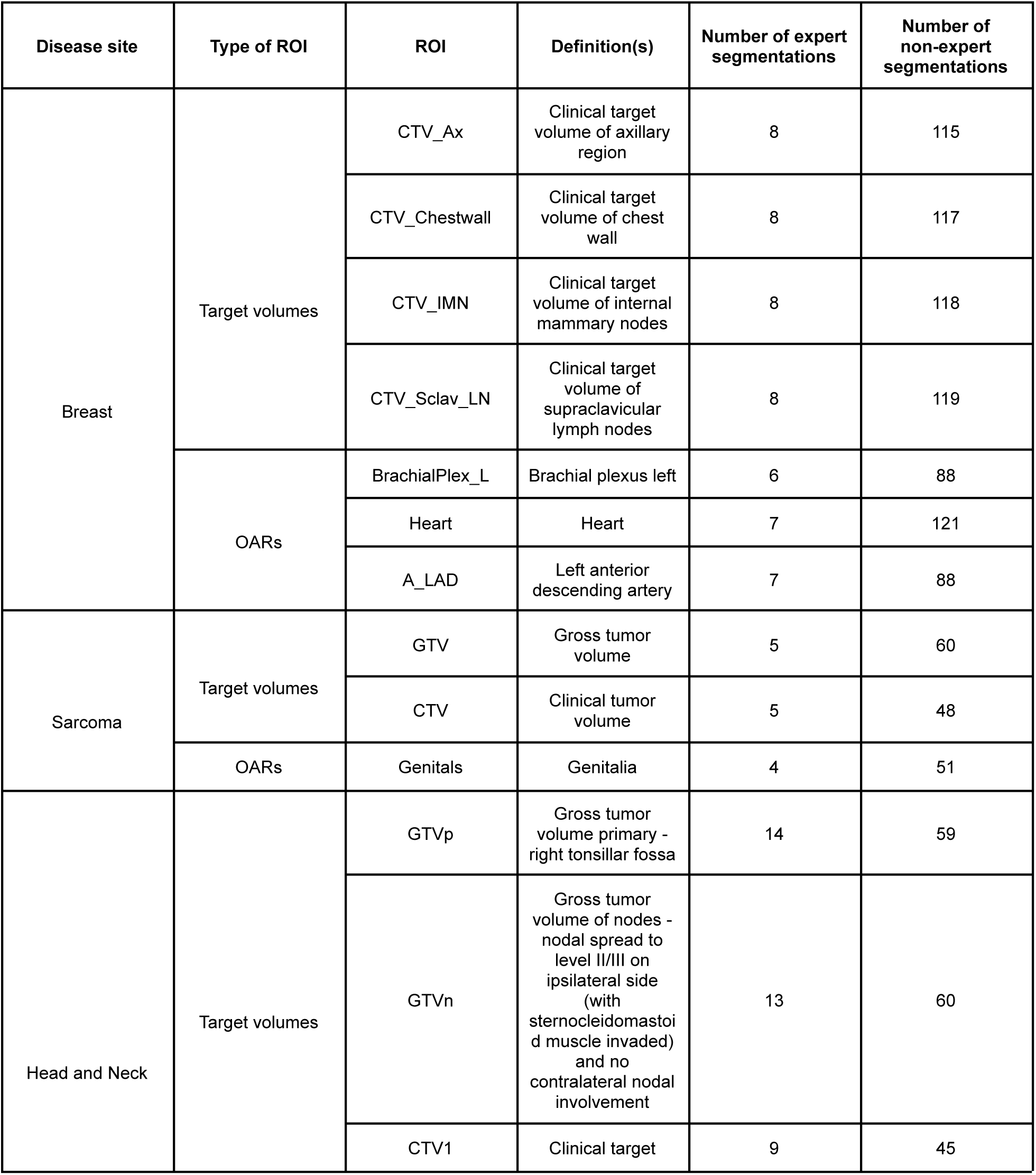
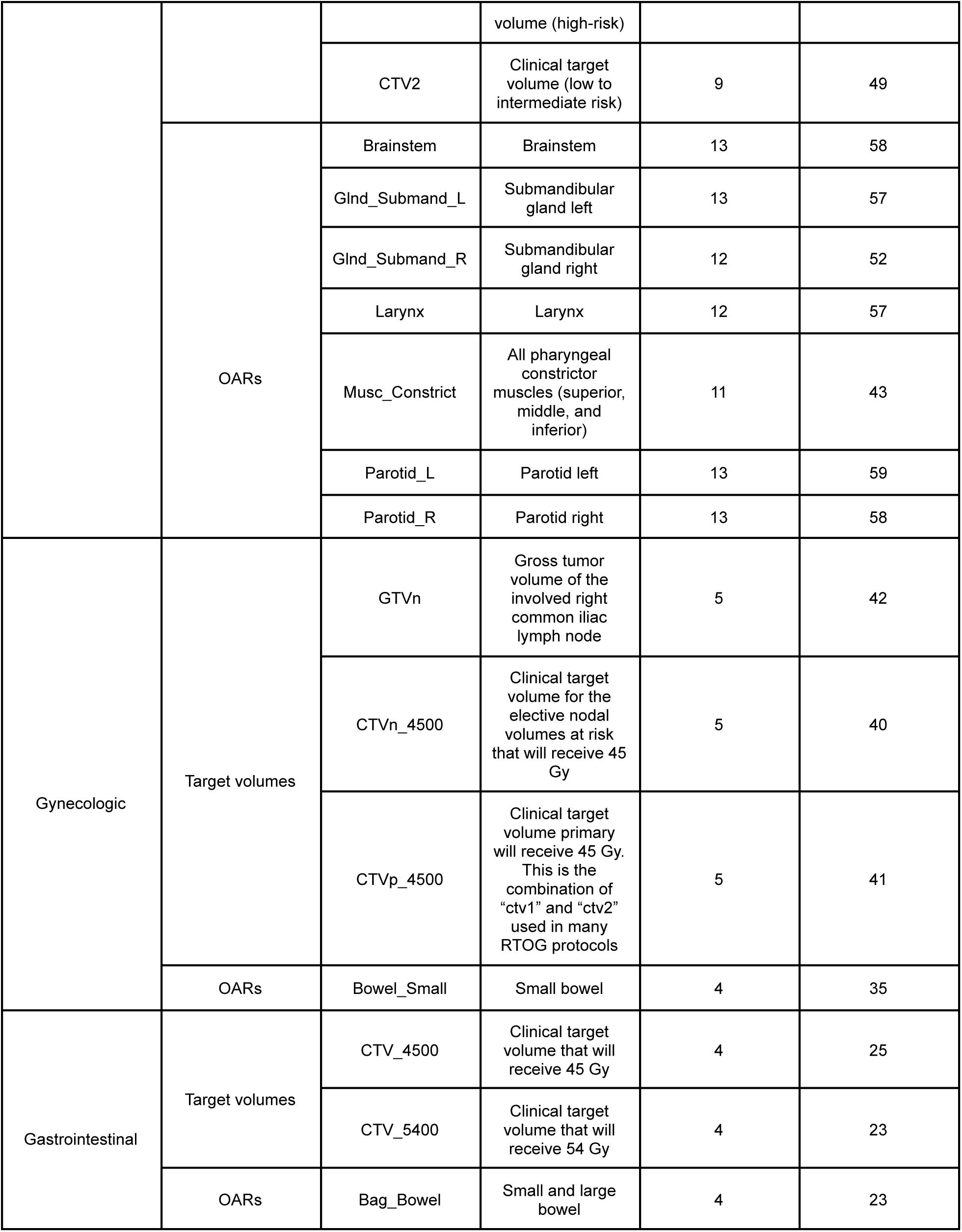
A complete list of the regions of interest (ROIs) used in this study for each disease site with the corresponding number of expert and non-expert segmentations available for each ROI. More information on these structures and the C3RO dataset as a whole can be found in the corresponding data descriptor (https://doi.org/10.1038/s41597-023-02062-w).

## Appendix B: Additional descriptive statistics and exploratory variable analysis

We calculated descriptive statistics for the radiation oncologist observers used in this study, including median and interquartile range values for numerical variables (total years of practice, number of colleagues) and percentages of binary categorical data (location, self-identified gender, practice type, self-identified race, treat site, academic affiliation, colleague presence). Values for each disease site were calculated separately, using observational data points at the region of interest level, which most accurately reflect the data pertinent to our analysis. Empty values for numerical values were ignored for these calculations. Descriptive statistics are shown in **Table B1**.

**Table B1.**
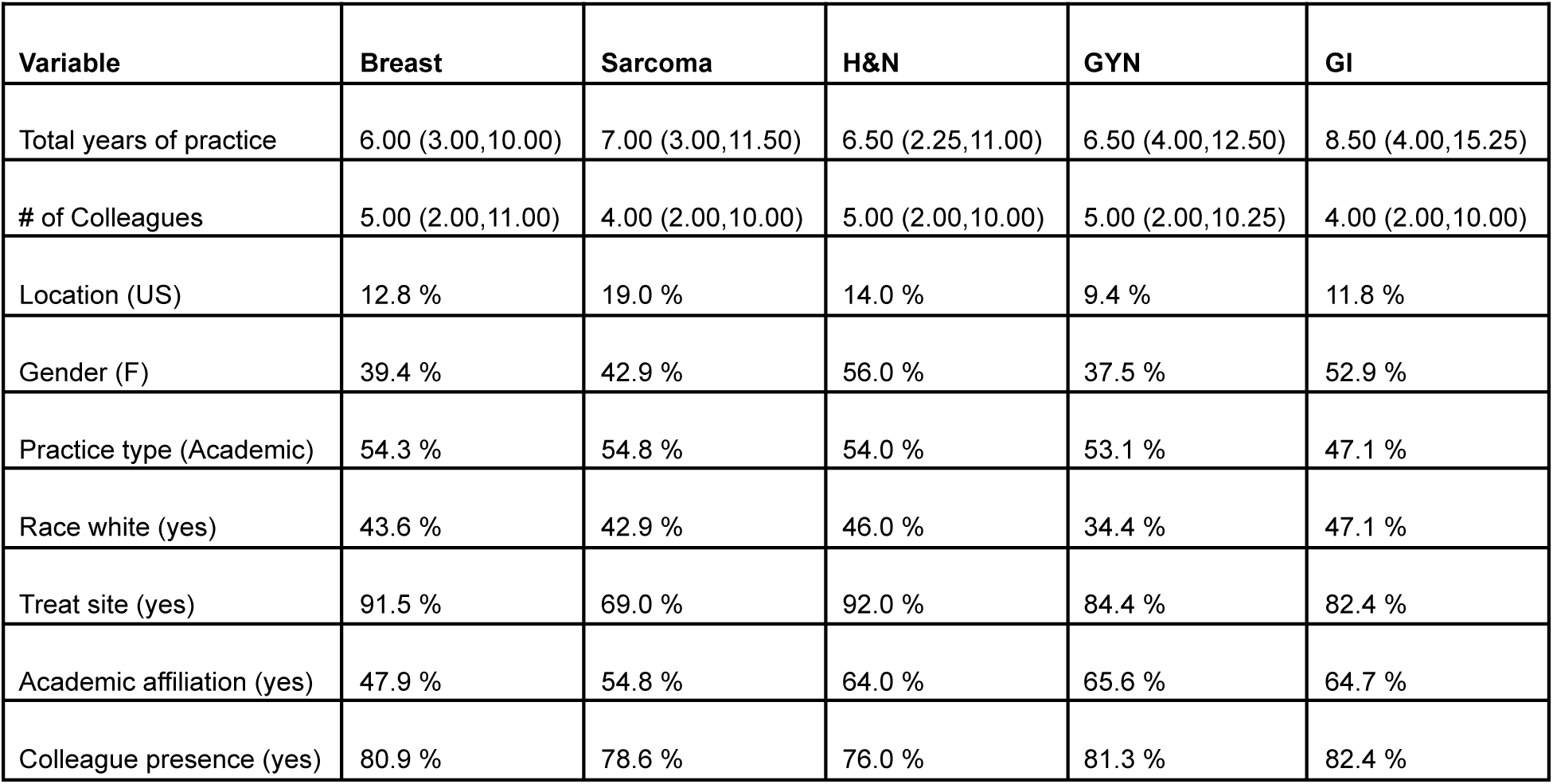
Descriptive statistics of demographic variables used in our analysis. Breast, sarcoma, head and neck (H&N), gynecologic (GYN), and gastrointestinal (GI) values were calculated separately. Median (interquartile range) values are shown for numerical variables. Percentages for a given binary class (indicated in parenthesis next to variable) are shown for the categorical variables.

| Variable | Breast | Sarcoma | H&N | GYN | GI |
| --- | --- | --- | --- | --- | --- |
| Total years of practice | 6.00 (3.00,10.00) | 7.00 (3.00,11.50) | 6.50 (2.25,11.00) | 6.50 (4.00,12.50) | 8.50 (4.00,15.25) |
| # of Colleagues | 5.00 (2.00,11.00) | 4.00 (2.00,10.00) | 5.00 (2.00,10.00) | 5.00 (2.00,10.25) | 4.00 (2.00,10.00) |
| Location (US) | 12.8 % | 19.0 % | 14.0 % | 9.4 % | 11.8 % |
| Gender (F) | 39.4 % | 42.9 % | 56.0 % | 37.5 % | 52.9 % |
| Practice type (Academic) | 54.3 % | 54.8 % | 54.0 % | 53.1 % | 47.1 % |
| Race white (yes) | 43.6 % | 42.9 % | 46.0 % | 34.4 % | 47.1 % |
| Treat site (yes) | 91.5 % | 69.0 % | 92.0 % | 84.4 % | 82.4 % |
| Academic affiliation (yes) | 47.9 % | 54.8 % | 64.0 % | 65.6 % | 64.7 % |
| Colleague presence (yes) | 80.9 % | 78.6 % | 76.0 % | 81.3 % | 82.4 % |

Using the demographic variables, we then performed an exploratory analysis to determine if any variables exhibited high correlations. A Spearman’s rank correlation was utilized since it could be utilized to analyze continuous numerical values and binary data simultaneously. Correlation heatmaps for each disease site are shown for each disease site in **Figures B1-B5**. After the exploratory analysis, academic affiliation was chosen to be excluded from the regression analysis due to its high correlation with practice type to enable greater model parsimony.

**Figure B1.**
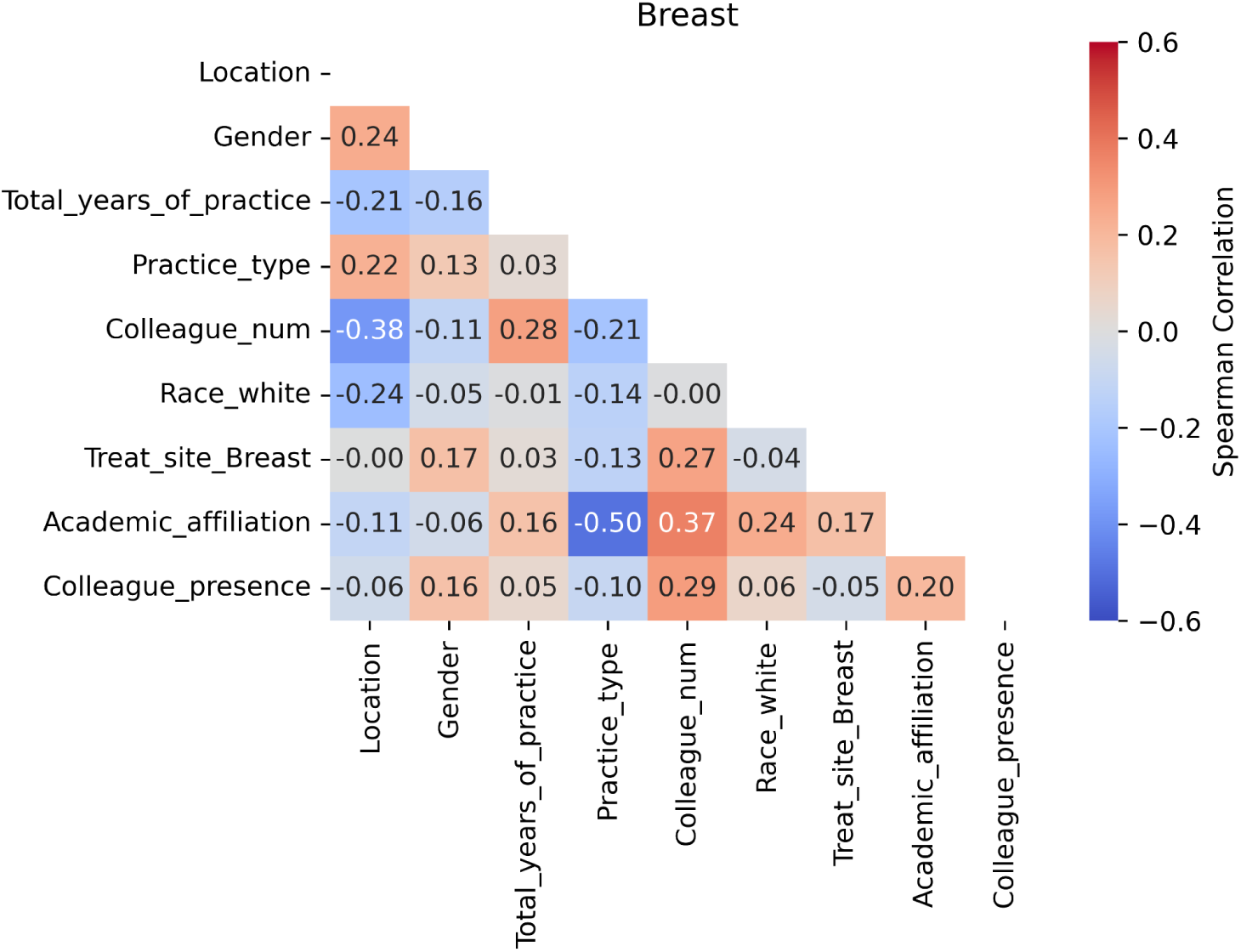
Correlation heatmap for the breast case.

**Figure B2.**
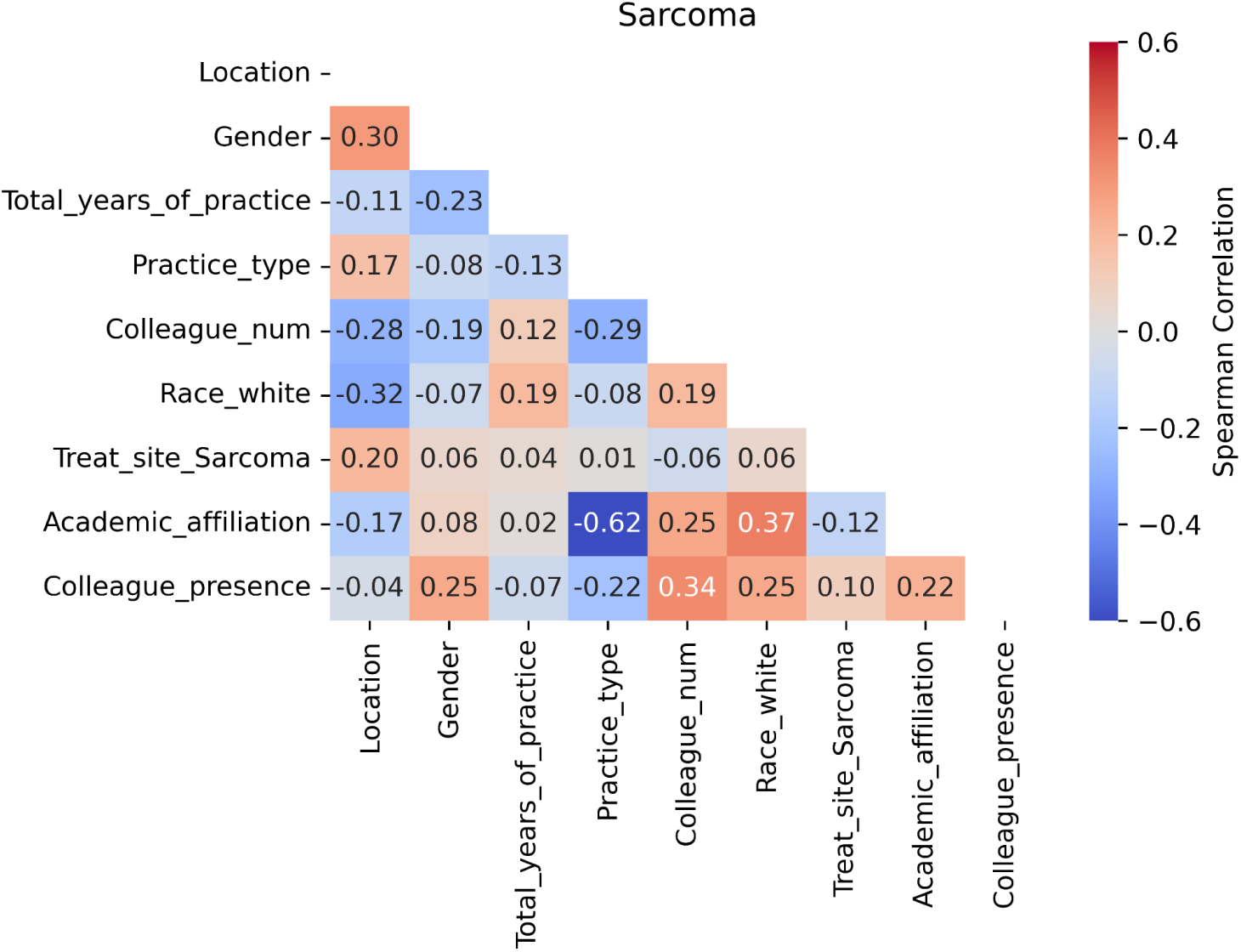
Correlation heatmap for the sarcoma case.

**Figure B3.**
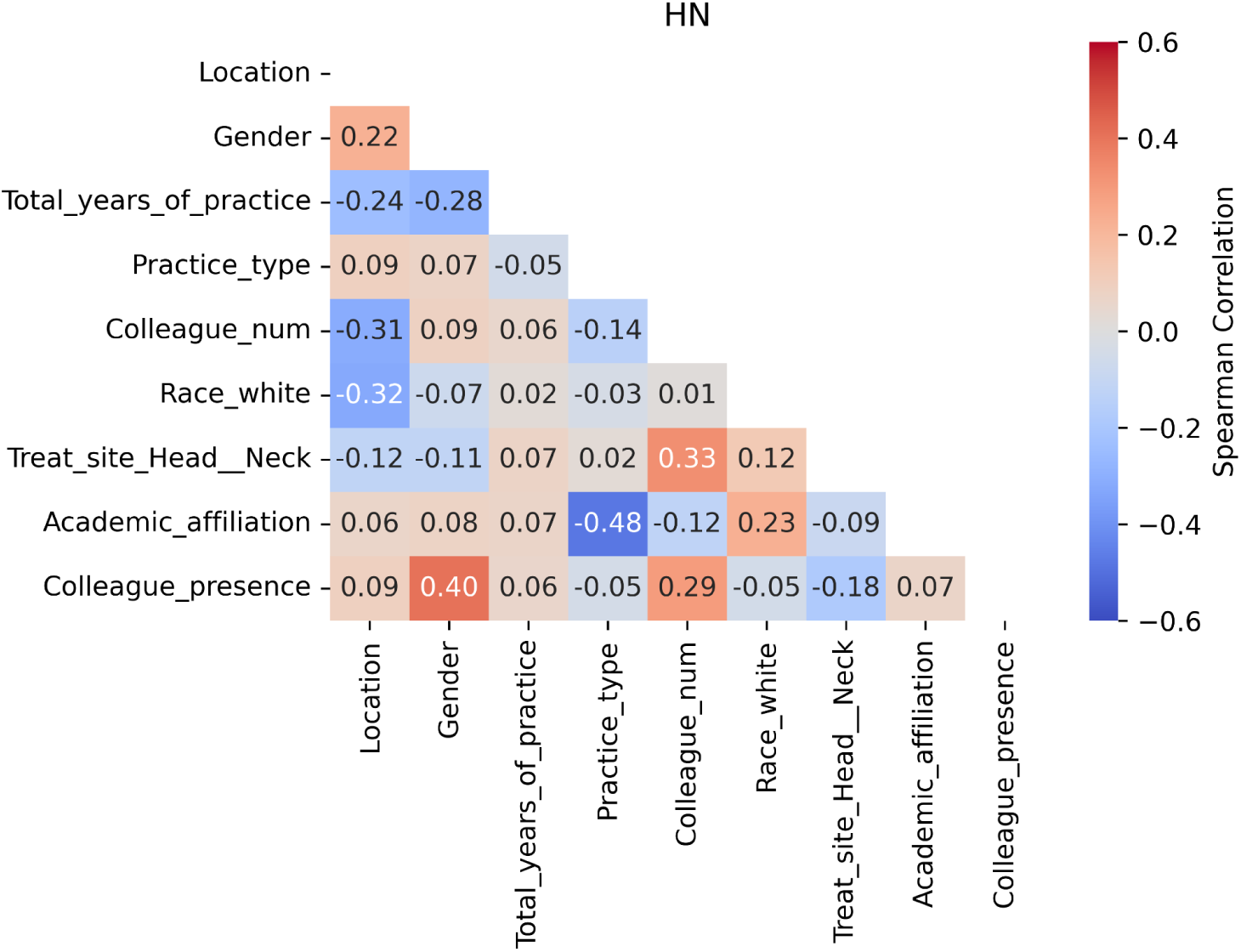
Correlation heatmap for the head and neck (HN) case.

**Figure B4.**
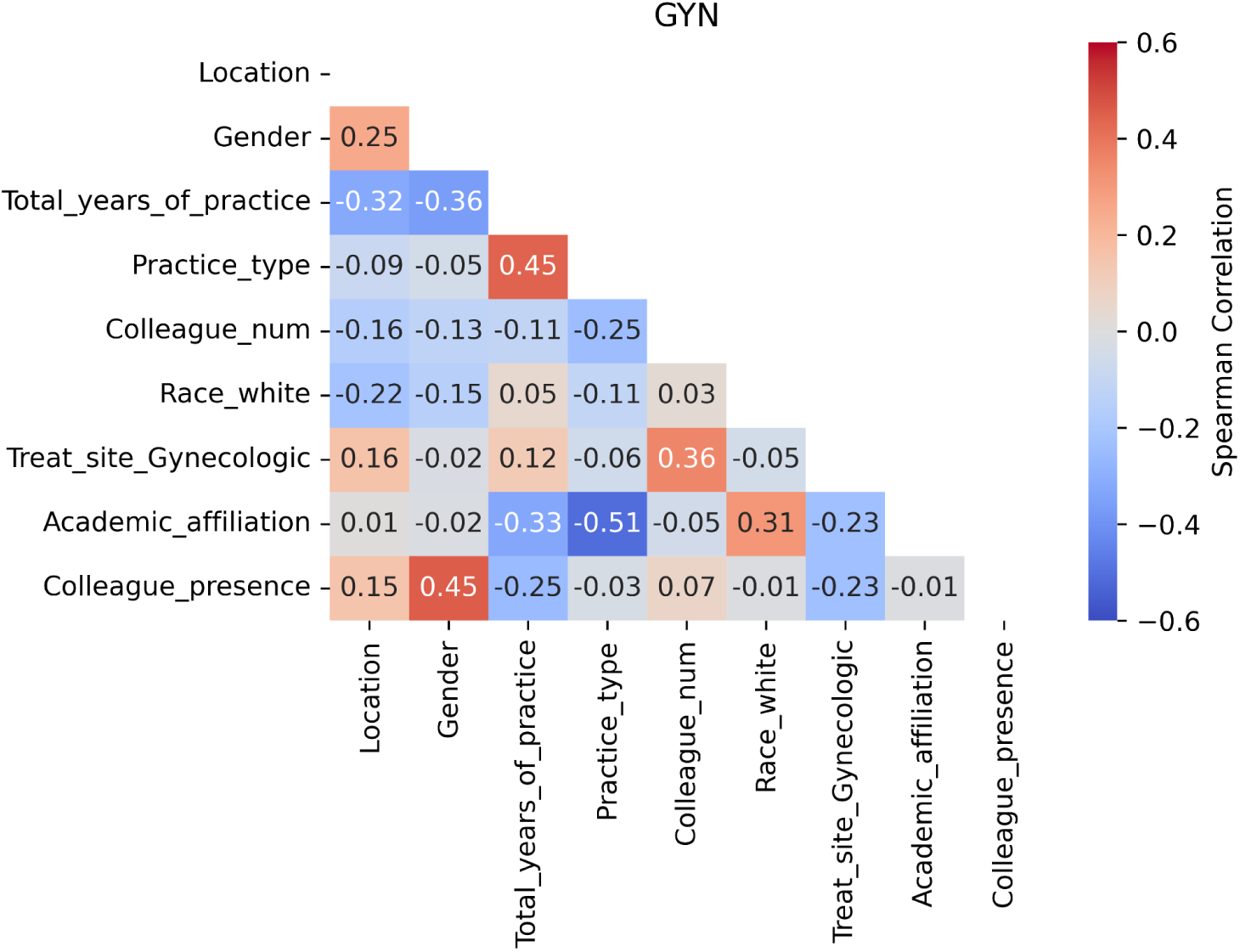
Correlation heatmap for the gynecologic (GYN) case.

**Figure B5.**
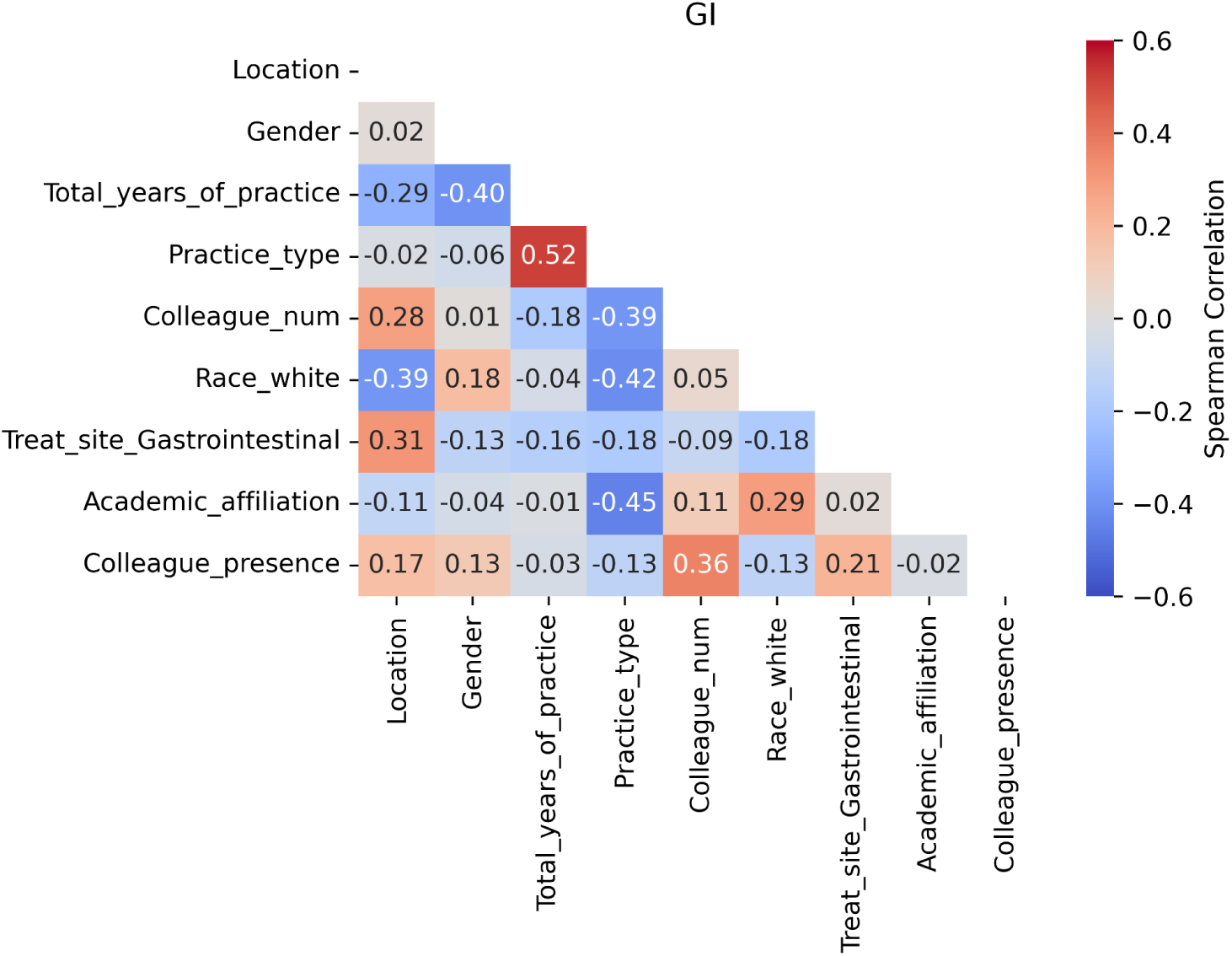
Correlation heatmap for the gastrointestinal (GI) case.

## Appendix C: Additional plots

**Figure C1.**
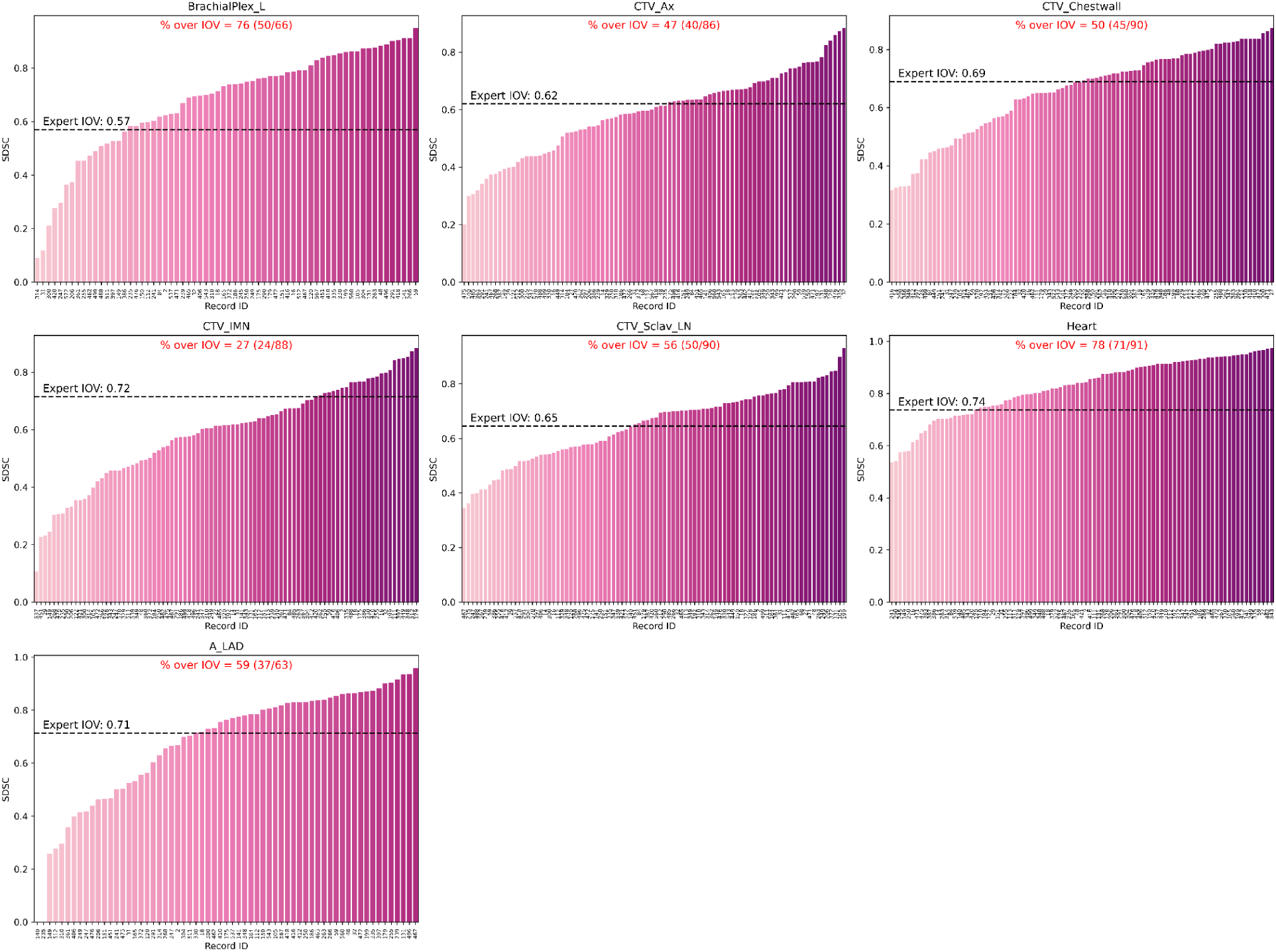
Barplots of individual observer segmentation performance vs. gold standard for the breast case using surface Dice similarity coefficient (SDSC). The gold standard segmentation is the consensus segmentation of all experts as derived from Simultaneous Truth and Performance Level Estimation (STAPLE). Black dotted lines indicate median expert interobserver SDSC for a corresponding region of interest. The percentage of observers that were able to cross the expert interobserver variability (IOV) cutoff are also shown in red above each plot.

**Figure C2.**
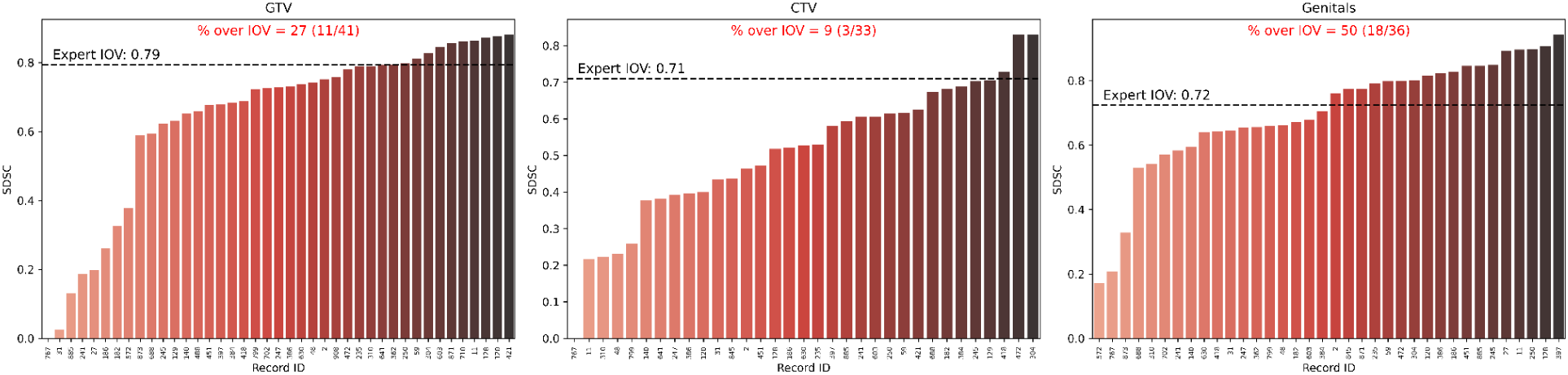
Barplots of individual observer segmentation performance vs. gold standard for sarcoma case using surface Dice similarity coefficient (SDSC). The gold standard segmentation is the consensus segmentation of all experts as derived from Simultaneous Truth and Performance Level Estimation (STAPLE). Black dotted lines indicate median expert interobserver SDSC for a corresponding region of interest. The percentage of observers that were able to cross the expert interobserver variability (IOV) cutoff are also shown in red above each plot.

**Figure C3.**
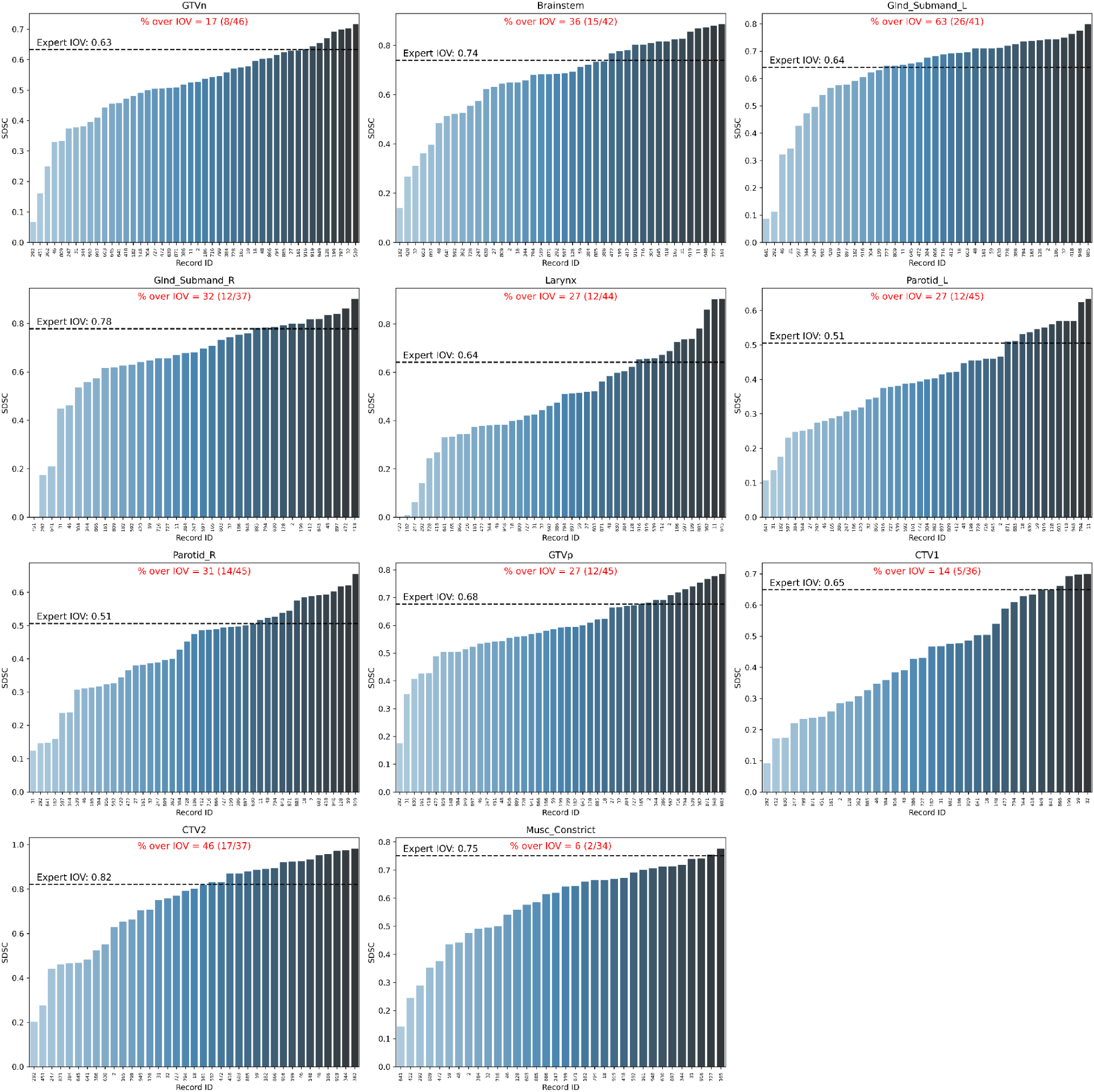
Barplots of individual observer segmentation performance vs. gold standard for head and neck case using surface Dice similarity coefficient (SDSC). The gold standard segmentation is the consensus segmentation of all experts as derived from Simultaneous Truth and Performance Level Estimation (STAPLE). Black dotted lines indicate median expert interobserver SDSC for a corresponding region of interest. The percentage of observers that were able to cross the expert interobserver variability (IOV) cutoff are also shown in red above each plot.

**Figure C4.**
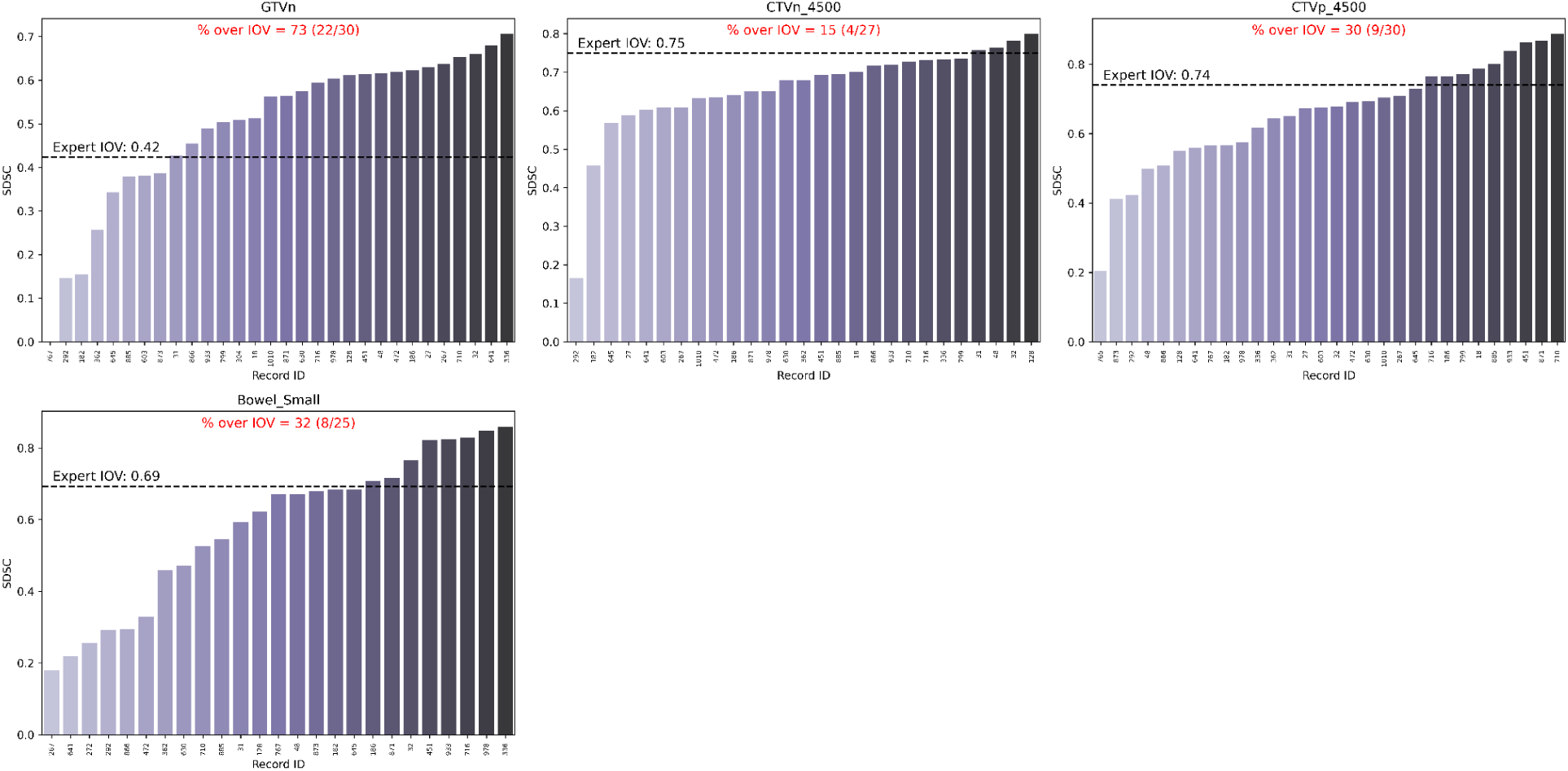
Barplots of individual observer segmentation performance vs. gold standard for gynecologic case using surface Dice similarity coefficient (SDSC). The gold standard segmentation is the consensus segmentation of all experts as derived from Simultaneous Truth and Performance Level Estimation (STAPLE). Black dotted lines indicate median expert interobserver SDSC for a corresponding region of interest. The percentage of observers that were able to cross the expert interobserver variability (IOV) cutoff are also shown in red above each plot.

**Figure C5.**
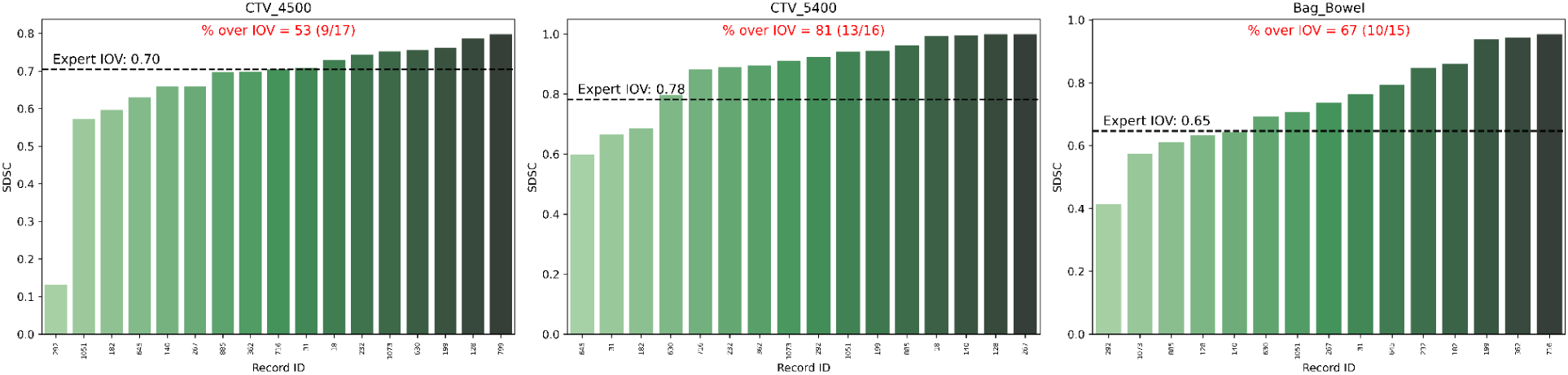
Barplots of individual observer segmentation performance vs. gold standard for gastrointestinal case using surface Dice similarity coefficient (SDSC). The gold standard segmentation is the consensus segmentation of all experts as derived from Simultaneous Truth and Performance Level Estimation (STAPLE). Black dotted lines indicate median expert interobserver SDSC for a corresponding region of interest. The percentage of observers that were able to cross the expert interobserver variability (IOV) cutoff are also shown in red above each plot.

## Appendix D: Additional information on Bayesian regression

Markov chain Monte Carlo convergence metric summary values were calculated for each model. **Tables D1-D10** display the Monte Carlo Standard Error of the mean (msce_mean), Monte Carlo Standard Error of the standard deviation (msce_sd), Effective Sample Size for the bulk of the distribution (ess_bulk), Effective Sample Size for the tail of the distribution (ess_tail), and the Gelman-Rubin statistic (r_hat) for each variable.

**Table D1.** Convergence parameters for the breast case using binarized DSC as the dependent variable.

| Case | Metric | Variable | mcse_mean | mcse_sd | ess_bulk | ess_tail | r_hat |
| --- | --- | --- | --- | --- | --- | --- | --- |
| Breast | DSC_binary | Intercept | 0.003 | 0.002 | 29568 | 22936 | 1 |
| Breast | DSC_binary | C(ROI_category)[Tumor] | 0.001 | 0.001 | 39951 | 24478 | 1 |
| Breast | DSC_binary | C(Location)[US] | 0.002 | 0.002 | 24293 | 20372 | 1 |
| Breast | DSC_binary | C(Gender, Treatment("Male"))[Female] | 0.002 | 0.001 | 25101 | 19633 | 1 |
| Breast | DSC_binary | Total_years_of_practice | 0.001 | 0.001 | 24629 | 19463 | 1 |
| Breast | DSC_binary | C(Practice_type, Treatment("Non-academic"))[Academic] | 0.001 | 0.001 | 25523 | 18624 | 1 |
| Breast | DSC_binary | Colleague_num | 0.001 | 0.001 | 27153 | 23344 | 1 |
| Breast | DSC_binary | C(Colleague_presence, Treatment("No"))[Yes] | 0.002 | 0.002 | 26875 | 21606 | 1 |
| Breast | DSC_binary | C(Race_white, Treatment("Unchecked"))[Checked] | 0.001 | 0.001 | 25907 | 18128 | 1 |
| Breast | DSC_binary | C(Treat_site_Breast, Treatment("Unchecked"))[Checked] | 0.003 | 0.002 | 26558 | 21617 | 1 |
| Breast | DSC_binary | 1\|Record_ID_sigma | 0.002 | 0.002 | 8348 | 10181 | 1 |

**Table D2.**
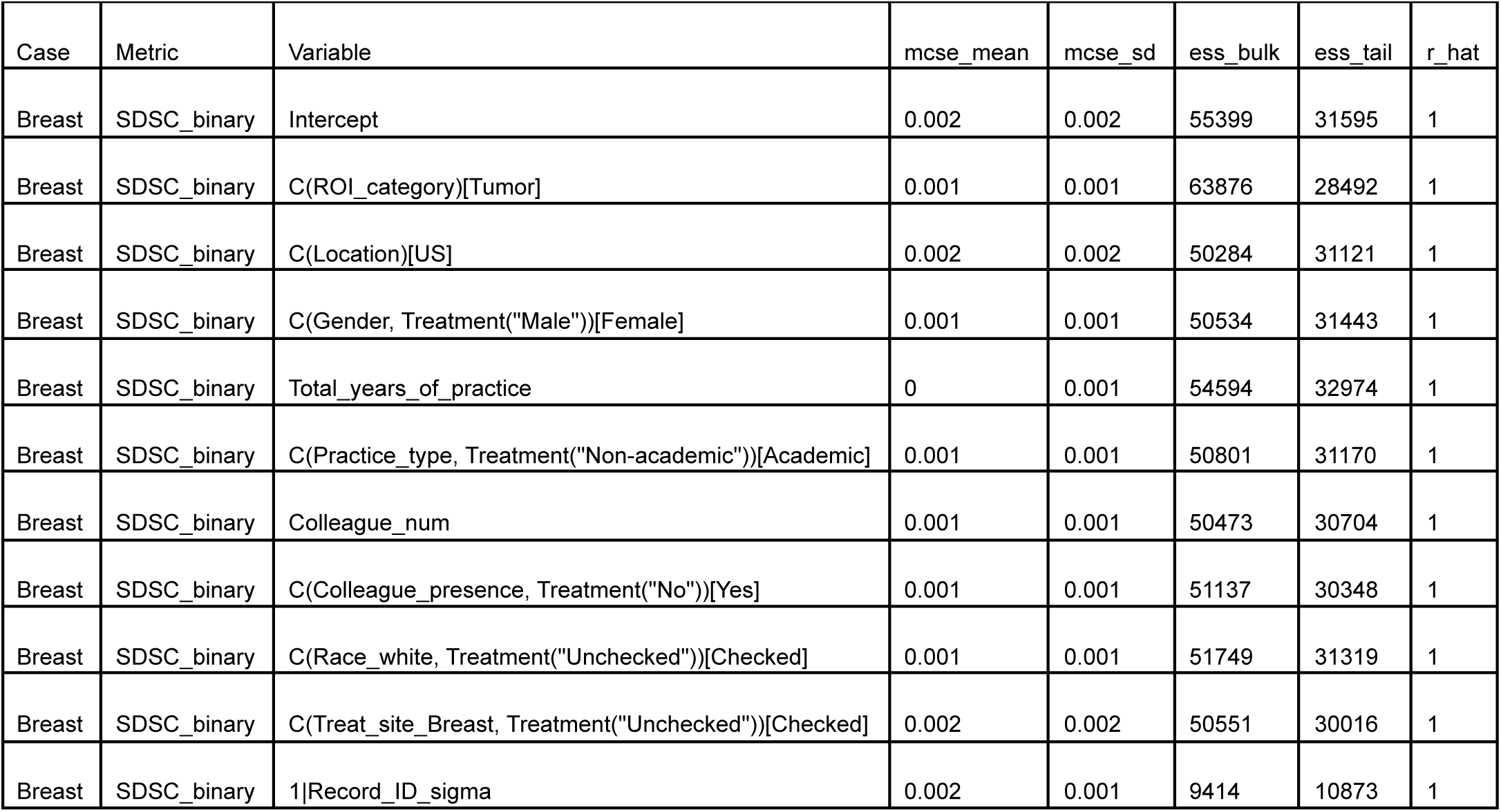
Convergence parameters for the breast case using binarized SDSC as the dependent variable.

**Table D3.** Convergence parameters for the sarcoma case using binarized DSC as the dependent variable.

| Case | Metric | Variable | mcse_mean | mcse_sd | ess_bulk | ess_tail | r_hat |
| --- | --- | --- | --- | --- | --- | --- | --- |
| Sarcoma | DSC_binary | Intercept | 0.013 | 0.012 | 12890 | 8384 | 1 |
| Sarcoma | DSC_binary | C(ROI_category)[Tumor] | 0.004 | 0.003 | 21729 | 17369 | 1 |
| Sarcoma | DSC_binary | C(Location)[US] | 0.009 | 0.007 | 14397 | 7301 | 1 |
| Sarcoma | DSC_binary | C(Gender, Treatment("Male"))[Female] | 0.006 | 0.006 | 16230 | 12533 | 1 |
| Sarcoma | DSC_binary | Total_years_of_practice | 0.008 | 0.01 | 5516 | 2137 | 1 |
| Sarcoma | DSC_binary | C(Practice_type, Treatment("Non-academic"))[Academic] | 0.013 | 0.018 | 5460 | 2422 | 1 |
| Sarcoma | DSC_binary | Colleague_num | 0.006 | 0.005 | 5777 | 2949 | 1 |
| Sarcoma | DSC_binary | C(Colleague_presence, Treatment("No"))[Yes] | 0.012 | 0.011 | 8407 | 4670 | 1 |
| Sarcoma | DSC_binary | C(Race_white, Treatment("Unchecked"))[Checked] | 0.011 | 0.009 | 6580 | 2933 | 1 |
| Sarcoma | DSC_binary | C(Treat_site_Sarcoma, Treatment("Unchecked"))[Checked] | 0.017 | 0.018 | 4992 | 2352 | 1 |
| Sarcoma | DSC_binary | 1\|Record_ID_sigma | 0.015 | 0.014 | 3533 | 1683 | 1 |

**Table D4.**
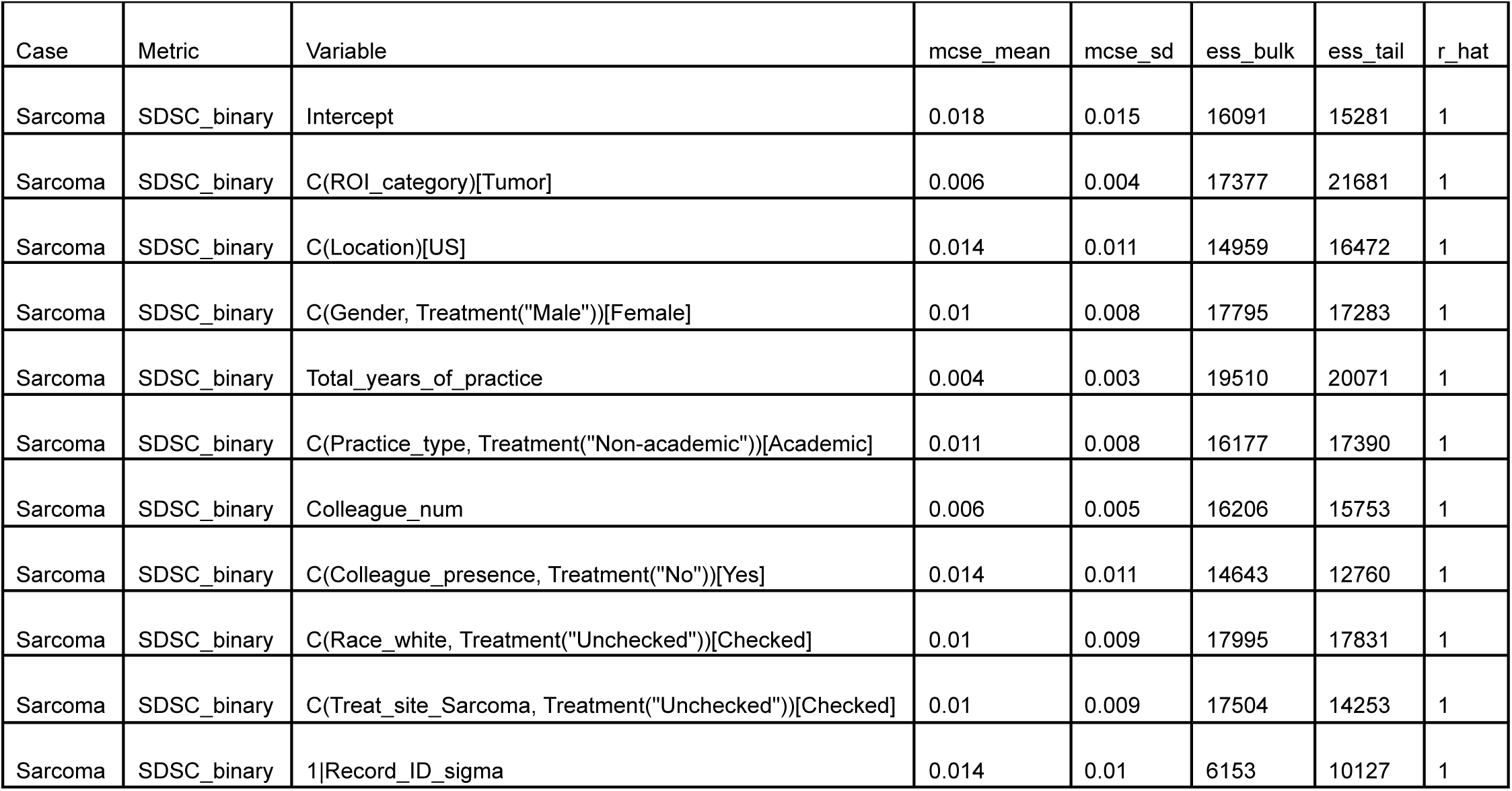
Convergence parameters for the sarcoma case using binarized SDSC as the dependent variable.

| Case | Metric | Variable | mcse_mean | mcse_sd | ess_bulk | ess_tail | r_hat |
| --- | --- | --- | --- | --- | --- | --- | --- |
| Sarcoma | SDSC_binary | Intercept | 0.018 | 0.015 | 16091 | 15281 | 1 |
| Sarcoma | SDSC_binary | C(ROI_category)[Tumor] | 0.006 | 0.004 | 17377 | 21681 | 1 |
| Sarcoma | SDSC_binary | C(Location)[US] | 0.014 | 0.011 | 14959 | 16472 | 1 |
| Sarcoma | SDSC_binary | C(Gender, Treatment("Male"))[Female] | 0.01 | 0.008 | 17795 | 17283 | 1 |
| Sarcoma | SDSC_binary | Total_years_of_practice | 0.004 | 0.003 | 19510 | 20071 | 1 |
| Sarcoma | SDSC_binary | C(Practice_type, Treatment("Non-academic"))[Academic] | 0.011 | 0.008 | 16177 | 17390 | 1 |
| Sarcoma | SDSC_binary | Colleague_num | 0.006 | 0.005 | 16206 | 15753 | 1 |
| Sarcoma | SDSC_binary | C(Colleague_presence, Treatment("No"))[Yes] | 0.014 | 0.011 | 14643 | 12760 | 1 |
| Sarcoma | SDSC_binary | C(Race_white, Treatment("Unchecked"))[Checked] | 0.01 | 0.009 | 17995 | 17831 | 1 |
| Sarcoma | SDSC_binary | C(Treat_site_Sarcoma, Treatment("Unchecked"))[Checked] | 0.01 | 0.009 | 17504 | 14253 | 1 |
| Sarcoma | SDSC_binary | 1\|Record_ID_sigma | 0.014 | 0.01 | 6153 | 10127 | 1 |

**Table D5.** Convergence parameters for the head and neck case using binarized DSC as the dependent variable.

| Case | Metric | Variable | mcse_mean | mcse_sd | ess_bulk | ess_tail | r_hat |
| --- | --- | --- | --- | --- | --- | --- | --- |
| HN | DSC_binary | Intercept | 0.004 | 0.004 | 37705 | 30054 | 1 |
| HN | DSC_binary | C(ROI_category)[Tumor] | 0.001 | 0.001 | 65671 | 30350 | 1 |
| HN | DSC_binary | C(Location)[US] | 0.004 | 0.003 | 31102 | 25877 | 1 |
| HN | DSC_binary | C(Gender, Treatment("Male"))[Female] | 0.003 | 0.002 | 33725 | 26508 | 1 |
| HN | DSC_binary | Total_years_of_practice | 0.001 | 0.001 | 34766 | 28060 | 1 |
| HN | DSC_binary | C(Practice_type, Treatment("Non-academic"))[Academic] | 0.002 | 0.002 | 35287 | 29790 | 1 |
| HN | DSC_binary | Colleague_num | 0.001 | 0.001 | 31819 | 26266 | 1 |
| HN | DSC_binary | C(Colleague_presence, Treatment("No"))[Yes] | 0.003 | 0.002 | 33815 | 28420 | 1 |
| HN | DSC_binary | C(Race_white, Treatment("Unchecked"))[Checked] | 0.002 | 0.002 | 32625 | 27214 | 1 |
| HN | DSC_binary | C(Treat_site_Head__Neck, Treatment("Unchecked"))[Checked] | 0.004 | 0.004 | 33875 | 26945 | 1 |
| HN | DSC_binary | 1\|Record_ID_sigma | 0.002 | 0.001 | 13010 | 19892 | 1 |

**Table D6.** Convergence parameters for the head and neck case using binarized SDSC as the dependent variable.

| Case | Metric | Variable | mcse_mean | mcse_sd | ess_bulk | ess_tail | r_hat |
| --- | --- | --- | --- | --- | --- | --- | --- |
| HN | SDSC_binary | Intercept | 0.005 | 0.004 | 28560 | 24200 | 1 |
| HN | SDSC_binary | C(ROI_category)[Tumor] | 0.001 | 0.001 | 50955 | 27532 | 1 |
| HN | SDSC_binary | C(Location)[US] | 0.004 | 0.003 | 23345 | 19662 | 1 |
| HN | SDSC_binary | C(Gender, Treatment("Male"))[Female] | 0.003 | 0.002 | 23809 | 20550 | 1 |
| HN | SDSC_binary | Total_years_of_practice | 0.001 | 0.001 | 25764 | 21664 | 1 |
| HN | SDSC_binary | C(Practice_type, Treatment("Non-academic"))[Academic] | 0.002 | 0.002 | 24808 | 21228 | 1 |
| HN | SDSC_binary | Colleague_num | 0.001 | 0.001 | 23480 | 21336 | 1 |
| HN | SDSC_binary | C(Colleague_presence, Treatment("No"))[Yes] | 0.003 | 0.003 | 22892 | 19482 | 1 |
| HN | SDSC_binary | C(Race_white, Treatment("Unchecked"))[Checked] | 0.002 | 0.002 | 25718 | 22428 | 1 |
| HN | SDSC_binary | C(Treat_site_Head__Neck, Treatment("Unchecked"))[Checked] | 0.004 | 0.004 | 26493 | 21941 | 1 |
| HN | SDSC_binary | 1\|Record_ID_sigma | 0.002 | 0.002 | 9903 | 15345 | 1 |

**Table D7.**
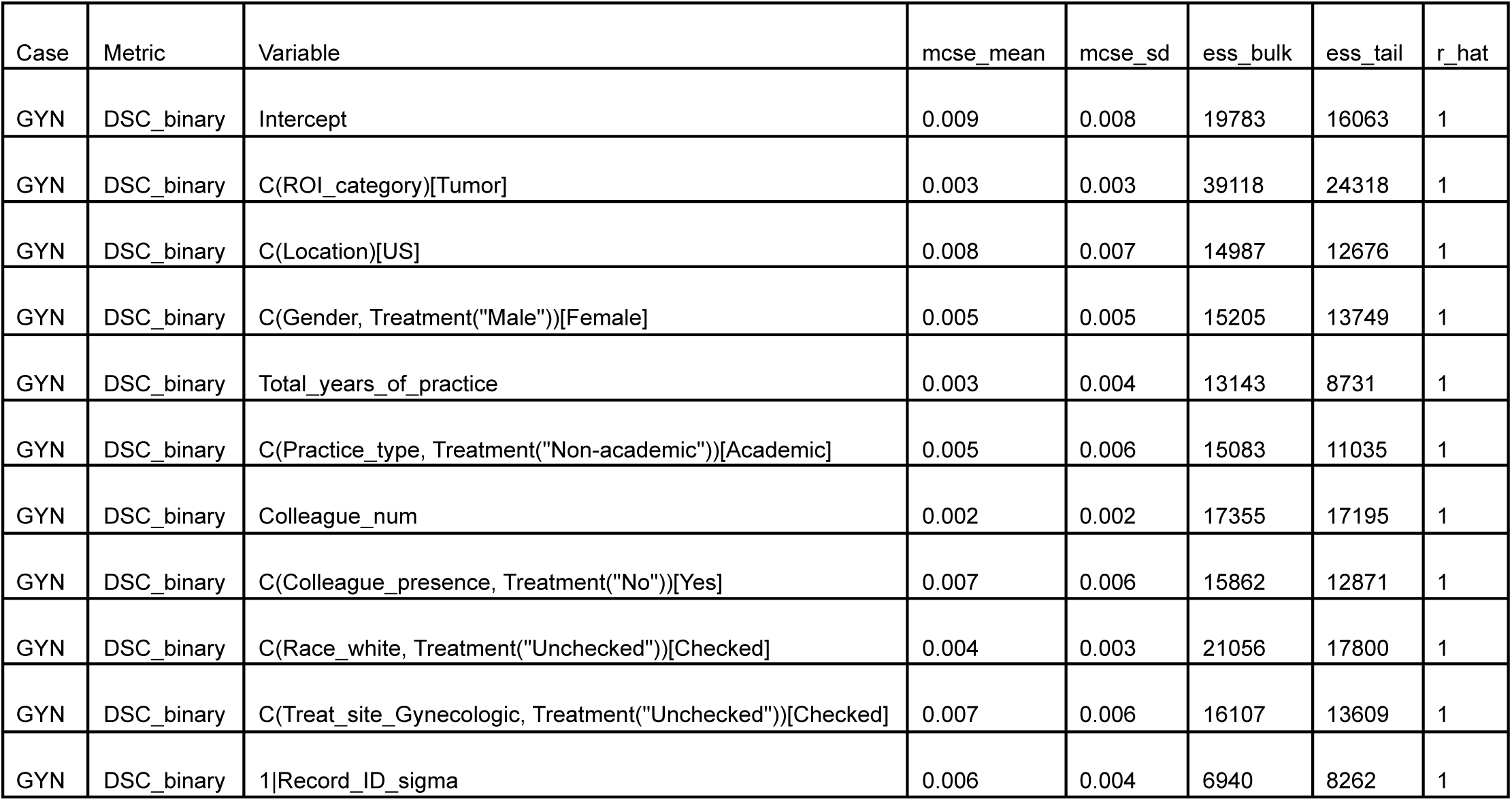
Convergence parameters for the gynecologic case using binarized DSC as the dependent variable.

| Case | Metric | Variable | mcse_mean | mcse_sd | ess_bulk | ess_tail | r_hat |
| --- | --- | --- | --- | --- | --- | --- | --- |
| GYN | DSC_binary | Intercept | 0.009 | 0.008 | 19783 | 16063 | 1 |
| GYN | DSC_binary | C(ROI_category)[Tumor] | 0.003 | 0.003 | 39118 | 24318 | 1 |
| GYN | DSC_binary | C(Location)[US] | 0.008 | 0.007 | 14987 | 12676 | 1 |
| GYN | DSC_binary | C(Gender, Treatment("Male"))[Female] | 0.005 | 0.005 | 15205 | 13749 | 1 |
| GYN | DSC_binary | Total_years_of_practice | 0.003 | 0.004 | 13143 | 8731 | 1 |
| GYN | DSC_binary | C(Practice_type, Treatment("Non-academic"))[Academic] | 0.005 | 0.006 | 15083 | 11035 | 1 |
| GYN | DSC_binary | Colleague_num | 0.002 | 0.002 | 17355 | 17195 | 1 |
| GYN | DSC_binary | C(Colleague_presence, Treatment("No"))[Yes] | 0.007 | 0.006 | 15862 | 12871 | 1 |
| GYN | DSC_binary | C(Race_white, Treatment("Unchecked"))[Checked] | 0.004 | 0.003 | 21056 | 17800 | 1 |
| GYN | DSC_binary | C(Treat_site_Gynecologic, Treatment("Unchecked"))[Checked] | 0.007 | 0.006 | 16107 | 13609 | 1 |
| GYN | DSC_binary | 1\|Record_ID_sigma | 0.006 | 0.004 | 6940 | 8262 | 1 |

**Table D8.** Convergence parameters for the gynecologic case using binarized SDSC as the dependent variable.

| Case | Metric | Variable | mcse_mean | mcse_sd | ess_bulk | ess_tail | r_hat |
| --- | --- | --- | --- | --- | --- | --- | --- |
| GYN | SDSC_binary | Intercept | 0.012 | 0.01 | 18014 | 12241 | 1 |
| GYN | SDSC_binary | C(ROI_category)[Tumor] | 0.003 | 0.004 | 31914 | 19393 | 1 |
| GYN | SDSC_binary | C(Location)[US] | 0.01 | 0.008 | 16197 | 15236 | 1 |
| GYN | SDSC_binary | C(Gender, Treatment("Male"))[Female] | 0.007 | 0.007 | 14827 | 12270 | 1 |
| GYN | SDSC_binary | Total_years_of_practice | 0.004 | 0.003 | 13708 | 9130 | 1 |
| GYN | SDSC_binary | C(Practice_type, Treatment("Non-academic"))[Academic] | 0.007 | 0.006 | 14715 | 10401 | 1 |
| GYN | SDSC_binary | Colleague_num | 0.004 | 0.003 | 13786 | 8826 | 1 |
| GYN | SDSC_binary | C(Colleague_presence, Treatment("No"))[Yes] | 0.009 | 0.008 | 13839 | 12240 | 1 |
| GYN | SDSC_binary | C(Race_white, Treatment("Unchecked"))[Checked] | 0.007 | 0.006 | 13981 | 10601 | 1 |
| GYN | SDSC_binary | C(Treat_site_Gynecologic, Treatment("Unchecked"))[Checked] | 0.009 | 0.008 | 14948 | 10891 | 1 |
| GYN | SDSC_binary | 1\|Record_ID_sigma | 0.008 | 0.006 | 5396 | 8252 | 1 |

**Table D9.** Convergence parameters for the gastrointestinal case using binarized DSC as the dependent variable.

| Case | Metric | Variable | mcse_mean | mcse_sd | ess_bulk | ess_tail | r_hat |
| --- | --- | --- | --- | --- | --- | --- | --- |
| GI | DSC_binary | Intercept | 0.015 | 0.013 | 32625 | 22474 | 1 |
| GI | DSC_binary | C(ROI_category)[Tumor] | 0.004 | 0.003 | 54623 | 28418 | 1 |
| GI | DSC_binary | C(Location)[US] | 0.02 | 0.016 | 26614 | 22297 | 1 |
| GI | DSC_binary | C(Gender, Treatment("Male"))[Female] | 0.007 | 0.007 | 28502 | 21852 | 1 |
| GI | DSC_binary | Total_years_of_practice | 0.005 | 0.004 | 20796 | 21685 | 1 |
| GI | DSC_binary | C(Practice_type, Treatment("Non-academic"))[Academic] | 0.011 | 0.009 | 20980 | 21353 | 1 |
| GI | DSC_binary | Colleague_num | 0.007 | 0.005 | 22249 | 23536 | 1 |
| GI | DSC_binary | C(Colleague_presence, Treatment("No"))[Yes] | 0.008 | 0.007 | 31745 | 22070 | 1 |
| GI | DSC_binary | C(Race_white, Treatment("Unchecked"))[Checked] | 0.007 | 0.006 | 31258 | 23044 | 1 |
| GI | DSC_binary | C(Treat_site_Gastrointestinal, Treatment("Unchecked"))[Checked] | 0.013 | 0.01 | 28288 | 24776 | 1 |
| GI | DSC_binary | 1\|Record_ID_sigma | 0.006 | 0.004 | 13999 | 15694 | 1 |

**Table D10.**
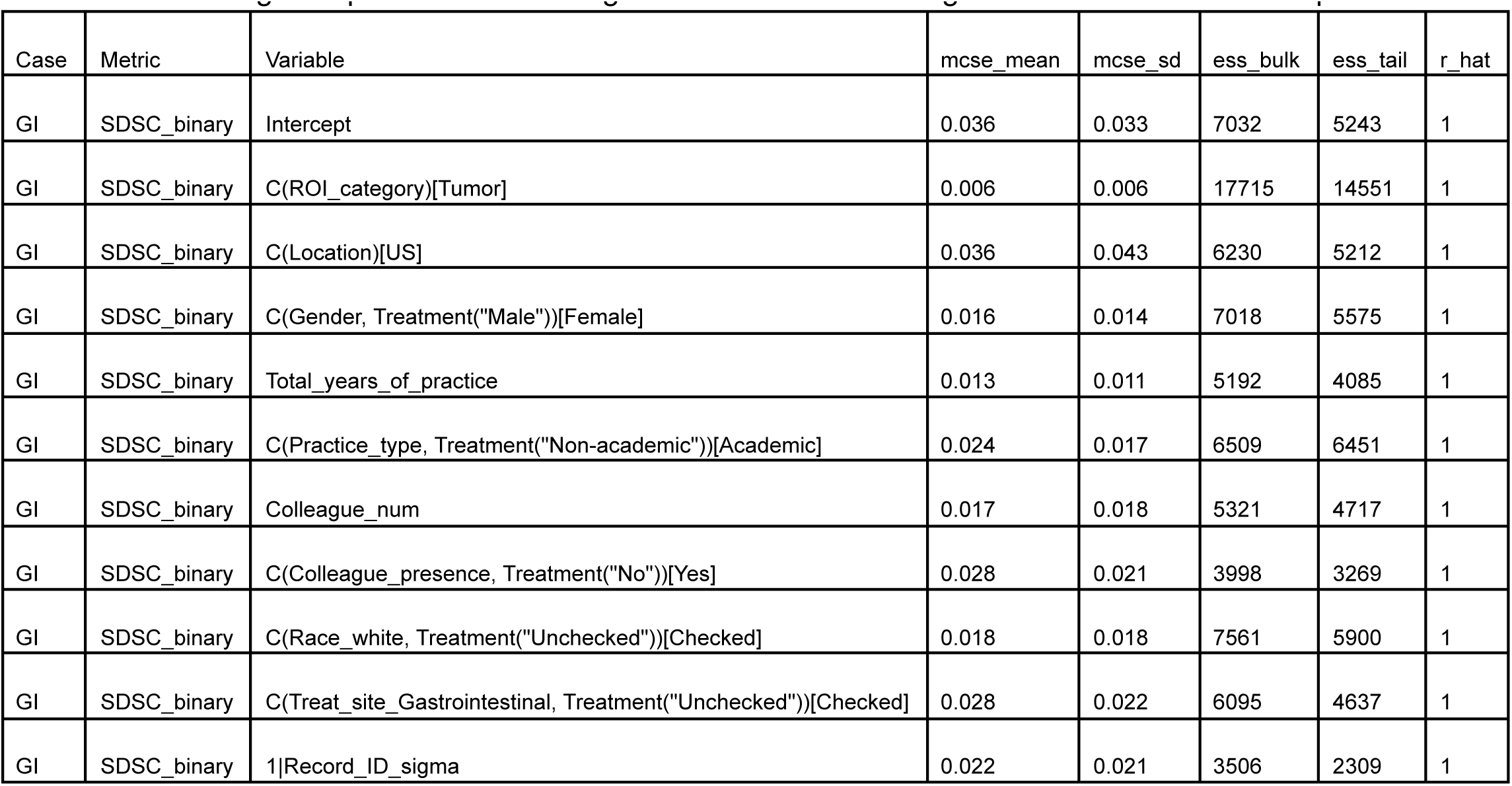
Convergence parameters for the gastrointestinal case using binarized SDSC as the dependent variable.

| Case | Metric | Variable | mcse_mean | mcse_sd | ess_bulk | ess_tail | r_hat |
| --- | --- | --- | --- | --- | --- | --- | --- |
| GI | SDSC_binary | Intercept | 0.036 | 0.033 | 7032 | 5243 | 1 |
| GI | SDSC_binary | C(ROI_category)[Tumor] | 0.006 | 0.006 | 17715 | 14551 | 1 |
| GI | SDSC_binary | C(Location)[US] | 0.036 | 0.043 | 6230 | 5212 | 1 |
| GI | SDSC_binary | C(Gender, Treatment("Male"))[Female] | 0.016 | 0.014 | 7018 | 5575 | 1 |
| GI | SDSC_binary | Total_years_of_practice | 0.013 | 0.011 | 5192 | 4085 | 1 |
| GI | SDSC_binary | C(Practice_type, Treatment("Non-academic"))[Academic] | 0.024 | 0.017 | 6509 | 6451 | 1 |
| GI | SDSC_binary | Colleague_num | 0.017 | 0.018 | 5321 | 4717 | 1 |
| GI | SDSC_binary | C(Colleague_presence, Treatment("No"))[Yes] | 0.028 | 0.021 | 3998 | 3269 | 1 |
| GI | SDSC_binary | C(Race_white, Treatment("Unchecked"))[Checked] | 0.018 | 0.018 | 7561 | 5900 | 1 |
| GI | SDSC_binary | C(Treat_site_Gastrointestinal, Treatment("Unchecked"))[Checked] | 0.028 | 0.022 | 6095 | 4637 | 1 |
| GI | SDSC_binary | 1\|Record_ID_sigma | 0.022 | 0.021 | 3506 | 2309 | 1 |

